# Genetics of Major Depressive Disorder in a Homogeneous Population with Uniform Phenotyping

**DOI:** 10.1101/2025.05.14.25325937

**Authors:** Floris Huider, Yuri Milaneschi, René Pool, Bernardo de A.P.C. Maciel, Scott D. Gordon, M. Liset Rietman, Almar A.L. Kok, Tessel E. Galesloot, Brittany L. Mitchell, Leen M. ‘t Hart, Femke Rutters, Marieke T. Blom, Didi Rhebergen, Marjolein Visser, Ingeborg A. Brouwer, Edith Feskens, Catharina A. Hartman, Albertine J. Oldehinkel, Mariska Bot, Eco J.C. de Geus, Lambertus A. Kiemeney, Martijn Huisman, H. Susan J. Picavet, W.M. Monique Verschuren, Nicholas G. Martin, Conor V. Dolan, Hanna M. van Loo, Brenda W.J.H. Penninx, Jouke-Jan Hottenga, Dorret I. Boomsma

## Abstract

Harmonized phenotyping and diverse population-specific studies are crucial for advancing gene discovery in psychiatric genetics. We conducted a genome-wide association (GWAS) mega-analysis of DSM-defined lifetime major depressive disorder (MDD) in 64 941 participants (25.7% cases) from the Dutch BIObanks Netherlands Internet Collaboration (BIONIC) consortium. SNP-based heritability was estimated at 13.4%, exceeding recent global meta-analyses, with a high genetic correlation (*r*_G_ = 0.89) to the latest major depression GWAS by the Psychiatric Genetics Consortium (PGC-MD). We identified a novel genome-wide significant locus in *PALMD* (*p* = 3.26 × 10⁻⁸), that was confirmed by GWAS-by-subtraction. Polygenic scores (PGSs) based on BIONIC predicted MDD in UK Biobank, and PGSs from PGC-MD predicted into BIONIC, with within-family analyses indicating minimal confounding. Genetic causal inference revealed associations with over 30 phenotypes. Twin concordance for MDD increased with polygenic burden, reinforcing its genetic architecture. This study emphasizes the power of harmonized phenotyping and regional biobanks in uncovering the genetic architecture of MDD, highlighting the value of population-specific studies for improving risk prediction and advancing psychiatric genetics.

## Introduction

Major depressive disorder (MDD) is among the most common psychiatric disorders with a lifetime prevalence that can reach up to 19% and is a leading cause of disability worldwide^1,2^. Episodes are frequently characterized by severe dysfunction and pervasiveness^3,4^. MDD is a genetically complex trait with a twin-based heritability of 37%^5^ and a polygenic architecture, characterized by the additive effect of many variants with small effects. In addition, MDD’s intrinsic heterogeneity, heterogeneity in depression diagnoses and assessment, and the relatively modest heritability compared to other psychiatric conditions have all been suggested as contributing to low statistical power for genome-wide association studies^6,7^. One successful approach to combating several of these problems is through large international collaborative efforts that bring together dozens of cohorts and millions of individuals^8–11^.

Concurrently, a discussion has emerged regarding the trade-off in sample size and phenotypic definition in genetic studies. Larger sample sizes have led to finding large numbers of genetic variants associated with broad concepts of depression. Growing sample sizes often come at the expense of uniform phenotyping; combining samples across multiple studies tends to yield a lower adherence to clinical diagnostic criteria and phenotypic uniformity. The use of self-reported diagnosis or treatment for depression, or depressed mood derived from a cut-off applied to a self-report symptom questionnaire in research, coined ‘minimal phenotyping’, may not align with MDD according to diagnostic criteria or represent associated but different genetic vulnerabilities^12–15^. In a comparison of depression phenotypes derived from the UK Biobank, Cai et al. (2020) found that minimal phenotyping definitions are epidemiologically distinct from strictly defined MDD, have lower SNP-based heritability (SNP-*h*²) estimates (11% for self-reported depression vs. 26% for MDD following diagnostic criteria), and that GWAS hits from minimal phenotyping may not be specific to MDD, and instead apply to a broader range of psychopathology and personality measures^15^. Consequently, follow-up characterization of GWAS loci from minimal phenotyping studies may not inform on biology specific to MDD.

The inclusion of data based on heterogeneous phenotypic assessment associates with a decay of SNP-*h*² estimates^16,17^. This has led to a call for approaches with uniform phenotyping, for example by adherence to clinical diagnostic criteria. In addition to phenotypic heterogeneity, meta-analyses can introduce genetic heterogeneity. Nearly all current studies tend to limit participation to participants of European ancestry, but even here there can be considerable population stratification and admixture affecting heterogeneity and GWAS results^18^, even after adjustments through ancestry-informative principal components. Tropf et al.^19^ find that heterogeneity across populations is one of the contributing factors to missing heritability, the common difference between GWAS-derived SNP-*h*² estimates and the upper limits set by twin-based approaches.

To address these challenges, we initiated the BIObanks Netherlands Internet Collaboration (BIONIC)^20^. This nationwide collaborative project combines genotype data from sixteen Dutch cohorts and focuses on uniform phenotyping of MDD according to clinical criteria. The Netherlands has a population of 18 million people, with ∼15.5 million of Dutch ancestry, and a population density of ∼535 people per square kilometer. In such single populations, population stratification is smaller, although not entirely absent^21,22^. We enriched the existing population and clinical cohorts in the Netherlands with an infrastructure for nationwide depression phenotyping so that all MDD cases were defined based on DSM-5 (Diagnostic and Statistical Manual of Mental Disorders)-criteria^3,23^. All cohorts agreed to share genotype and phenotype data in a central location for mega-analysis, where QC and genotype imputation were conducted simultaneously and harmonized across all arrays and participants.

With the BIONIC dataset, we ask the question if we can find evidence for a contribution of single nucleotide polymorphisms (SNP) to MDD as evidenced by genome-wide significant associations, significant SNP heritability, and polygenic prediction. We report results from the mega-analysis of lifetime MDD case-control status and genome-wide SNP data in 64 941 individuals (N_case_ = 16 655, N_control_ = 48 286). We leverage clinical diagnostic criteria in a relatively homogeneous sample to identify genetic variant associations and consider the benefits of such an approach for capturing genetic signal, comparing our results to the largest GWAS of major depression to date (by the Psychiatric Genetics Consortium MDD workgroup)^10^. We apply GWAS-by-subtraction, in which we searched for a uniquely Dutch MDD signal by ‘subtracting’ the largest global major depression GWAS effort from our GWAS results. We extend the GWA mega-analysis of MDD with a series of complementary analyses. These include fine-mapping approaches, such as gene-based tests and tissue expression analyses, as well as broader investigations into phenome-wide genetic correlations and causal relationships. We evaluate polygenic risk scores (PGS) through in- and out-of-sample predictions, assess potential confounding via within- and between-family tests, and examine twin concordance across PGS deciles.

## Methods

### Sample and phenotype descriptions

This genome-wide association (GWAS) mega-analysis combines lifetime major depressive disorder (MDD) and genotype data from sixteen Dutch cohorts in the BIONIC (BIObanks Netherlands Internet Collaboration) project. Descriptions of the sixteen cohorts can be found in the Supplementary Information. The majority of phenotype data were collected through the Lifetime Depression Assessment of Survey (LIDAS)^23^, developed to assess DSM-5 MDD. Other data were derived from clinical interviews and questionnaires (see eMethods for a detailed description). MDD cases were uniformly defined according to DSM-5 diagnostic criteria as ever having had a period of at least two weeks with five or more depression symptoms causing dysfunction in life, of which at least one a cardinal symptom^24^. MDD controls were defined as having no such period, and – when information on other psychopathology was available – having no other psychopathology (eMethods). The final analysis set included 64 941 European ancestry individuals with non-missing MDD, covariate (sex, age), and genotype data;16 655 lifetime MDD cases and 48 286 screened controls.

### Genotyping, quality control, and imputation

Details on the quality control (QC) procedure have been described previously^20^. In brief, the sixteen cohorts had collected genotype data on seven different arrays across multiple batches, resulting in nineteen datasets. This included some smaller datasets that could not be analyzed independently because they were too small or had an overrepresentation of cases. To include the largest number of individuals, SNP data genotyped on the same arrays were combined after basic sample and SNP quality control (details in eMethods). The resulting array groups were the units for further QC and imputation. Genotype data were imputed against the Human Reference Consortium (HRC) panel (v1.1)^25^. After imputation, the array groups had identical genome coverage and were merged into a single set for mega-analysis. All arrays included genotype data on the 22 autosomes. The availability of X-chromosome data varied (Supplementary Figure 1), with the first pseudo-autosomal region (PAR), non-PAR, and second PAR available in 55 141, 55 863, and 41 441 individuals, respectively.

### Genome-wide association mega-analysis

We conducted a GWAS mega-analysis of lifetime MDD for SNPs on the merged BIONIC dataset. Analyses were conducted in fastGWA^26^ from the Genome-wide Complex Trait Analysis (GCTA) software^27^, with a generalized linear mixed model. SNPs with minor allele frequency < 0.01 and imputation quality < 0.40 were excluded from analysis. Analyses were corrected for sex, age, 10 ancestry-informative principal components (PCs), genotype array, and relatedness (through a genetic relationship matrix). Independent GWAS signals were identified by conditional and joint analysis in GCTA^28^, where the BIONIC GWAS MDD sample served as the LD reference sample, restricted to one individual per related pair (relatedness ≥ 0.125) as calculated by the KING software (V2.2.6)^29^.

### SNP-based heritability and genetic correlation

SNP-based heritability (SNP-*h*²) and genetic correlation were estimated by Linkage Disequilibrium score regression (v1.0.1; LDSC)^30^. Effective N was calculated following previous literature^31^. SNP-*h²* was estimated on the liability scale, specifying 18% as population prevalence^3^ and sample prevalence as observed in the data (27% for the main GWAS model).

### GWAS-by-subtraction

To explore if there was evidence for specific Dutch MDD genetic signal, we carried out GWAS-by-subtraction as implemented in the genomic structural equation modeling (Genomic SEM)^32^ software with the European GWAS summary statistics from the largest major depression (MD) GWAS to date^10^ (PGC-MD, excluding Dutch samples) and the BIONIC MDD GWAS as input. GWAS-by-subtraction performs a GWAS of the unique genetic variation in BIONIC MDD GWAS after regressing out the variance shared with the PGC-MD GWAS, i.e., the variance not shared between the two traits^33^.

### Polygenic score prediction

We explored genetic risk prediction of lifetime MDD through in-sample and out-of-sample polygenic score (PGS) prediction with PGSs generated in the LDpred software (v0.9.1)^34^. For in-sample prediction, we generated MD PGSs in BIONIC based on the PGC-MD GWAS summary statistics (European samples and including 23andMe), excluding all datasets from the Netherlands^10^. The association between the MD PGS and lifetime MDD case-control status in BIONIC was estimated by logistic regression in unrelated individuals with age at assessment, sex, and 10 ancestry informative PCs as covariates. The proportion of variance explained by the PGS on the liability scale for MDD case-control status was estimated according to Lee et al.^35^.

We explored out-of-sample MD PGS prediction based on the BIONIC MDD GWAS summary statistics in the UK Biobank, for a strict definition of MDD (clinical criteria) and broad depression (clinical criteria, sumscore cut-offs, or self-report single item measures). Details on data field codes and genotype QC are in eMethods. A total of 23 755 and 132 122 individuals met the criteria for strict and broad phenotype definitions, respectively. PGS were generated similarly to the in-sample prediction in LDpred (v1.0.10) and PLINK (v1.9)^36^ assuming an infinitesimal model. PGS prediction was estimated by logistic regression in unrelated individuals with age at measurement, sex, genotype array, and 10 PCs as covariates.

Within-family designs can account for many sources of passive genotype-environment correlation, with dizygotic (DZ) twins having the additional benefit that all shared environmental factors are time-invariant among twins^37^. We sought to evaluate MD PGS prediction in a within-family design, following the approach of Selzam et al.^37^ in 1141 dizygotic twins with non-missing data (lifetime MDD and PGS) from the Netherlands Twin Register (NTR; a collaborating cohort in BIONIC). First, PGS prediction of MDD was assessed in the entire DZ twin sample through a logistic generalized linear mixed-effects model, correcting for sex and genotype array. Next, between-family PGS effects were defined as the change in lifetime MDD risk with a change in mean family PGS values, and within-family PGS effects as the change in lifetime MDD risk with a change in the difference between individual PGS and the family average PGS. The two were fit as predictors of MDD in the same model, together with a random effect for family, so that individual estimates are adjusted for and independent of the effect of the other estimate. The statistical difference between the between- and within-family PGS effects were then empirically tested through a chi-square test of the difference between their coefficients divided by the standard deviations of the sampling distribution of the estimate differences^38,39^.

Following a recent example in bipolar disorder and schizophrenia^40^, we computed MD PGS deciles in N = 2963 complete monozygotic and dizygotic twin pairs from the NTR cohort who took part in BIONIC. We then distinguished three groups: concordantly affected (133) pairs, concordantly unaffected (2257) pairs, and discordant (573) pairs. We hypothesized that concordantly affected twins would be overrepresented in high MD PGS deciles. We repeated this approach in an independent sample for replication, the Australian Genetics of Depression Study^41^ (Supplementary Information). We formally tested the association between the mean MD PGS in NTR twin pairs and MDD concordance in an ordinal logistic regression model, correcting for two ancestry-informative PCs^40^.

### Genetic causal associations

We computed genetic correlations between the BIONIC MDD GWAS and 1461 disease, personality, and lifestyle traits using bivariate LDSC in the Complex-Traits Genetics Virtual Lab (CTG-VL) analysis pipeline (https://vl.genoma.io/). Details on their derivation and acquisition are given in the eMethods. We then applied the bivariate latent causal variable (LCV) model to traits with a significant genetic correlation with MDD. The LCV method identifies potentially causal relationships among heritable traits^42^, quantifying the degree to which a genetic correlation between two traits may be explained by (partial) genetic causality as opposed to full horizontal pleiotropy through the genetic causal proportion (GCP) metric. Benjamini-Hochberg’s False Discovery Rate < 5% was applied to correct for multiple testing at both the genetic correlation and LCV steps.

### Gene-level analyses & tissue enrichment

We performed a gene-based test and gene-set analysis of MDD in the MAGMA (Multi-marker Analysis of GenoMic Annotation; v1.08)^43^ tool, mapping SNPs from the BIONIC MDD GWAS to 19 210 protein-coding genes (eMethods). Gene sets were derived from categories C2 and C5 from the publicly-accessible MsigDB v7.0. Significance was set at the Bonferroni-corrected threshold of *p* = 0.05 / 15 488 = 3.23 × 10^-6^. We further performed tissue expression analysis for 53 tissue categories in MAGMA with the gene-based test results as input. Gene expression datasets were obtained from GTEx v8 RNAseq. Significance was defined as *p* = 0.05 / 18 062 = 2.77 × 10^-6^.

### Gene prioritization

We fine-mapped significant loci in the BIONIC MDD GWAS in the Fine-mapped Locus Assessment Model of Effector geneS (FLAMES)^44^ software, to predict the most likely effector gene in a locus (eMethods).

### Sensitivity analyses

We ran several sensitivity analyses to assess the robustness of outcomes and the validity of the approach (eMethods). First, we ran a GWAS mega-analysis of height in the BIONIC data to benchmark the MDD results. Second, we ran a GWAS meta-analysis of lifetime MDD to compare against the mega-analysis approach. Third, we repeated the GWAS mega-analysis for an alternative lifetime MDD phenotype definition in which controls were screened less strictly.

## Results

### Genome-wide association mega-analysis of lifetime MDD

We ran a genome-wide association (GWAS) mega-analysis of lifetime major depressive disorder (MDD) in 64 941 Dutch individuals (16 655 lifetime MDD cases and 48 286 screened controls) (Figure 1, Supplementary Figs. 2, 3). The majority of the sample was female (60.8% in the full sample) and the average age was 50.7 (SD 16.0) (Supplementary Table 1). The rs3818852 SNP on chromosome 1, an intronic variant of the *PALMD* gene, reached significance at the genome-wide-adjusted alpha level (OR = 0.93, *p* = 3.26 × 10^-8^). *PALMD* encodes the palmdelphin protein, involved in cellular processes including cytoskeletal organization and cell shape modulation. There have been no previous GWAS hits for rs3818852 registered in the GWAS catalog, indicating a potentially novel cross-domain phenotypic association. Supplementary Table 2 lists the 10 most significant independent SNPs, with the summary statistics for the top 1000 loci available in the Supplementary Material.

**Figure 1.**
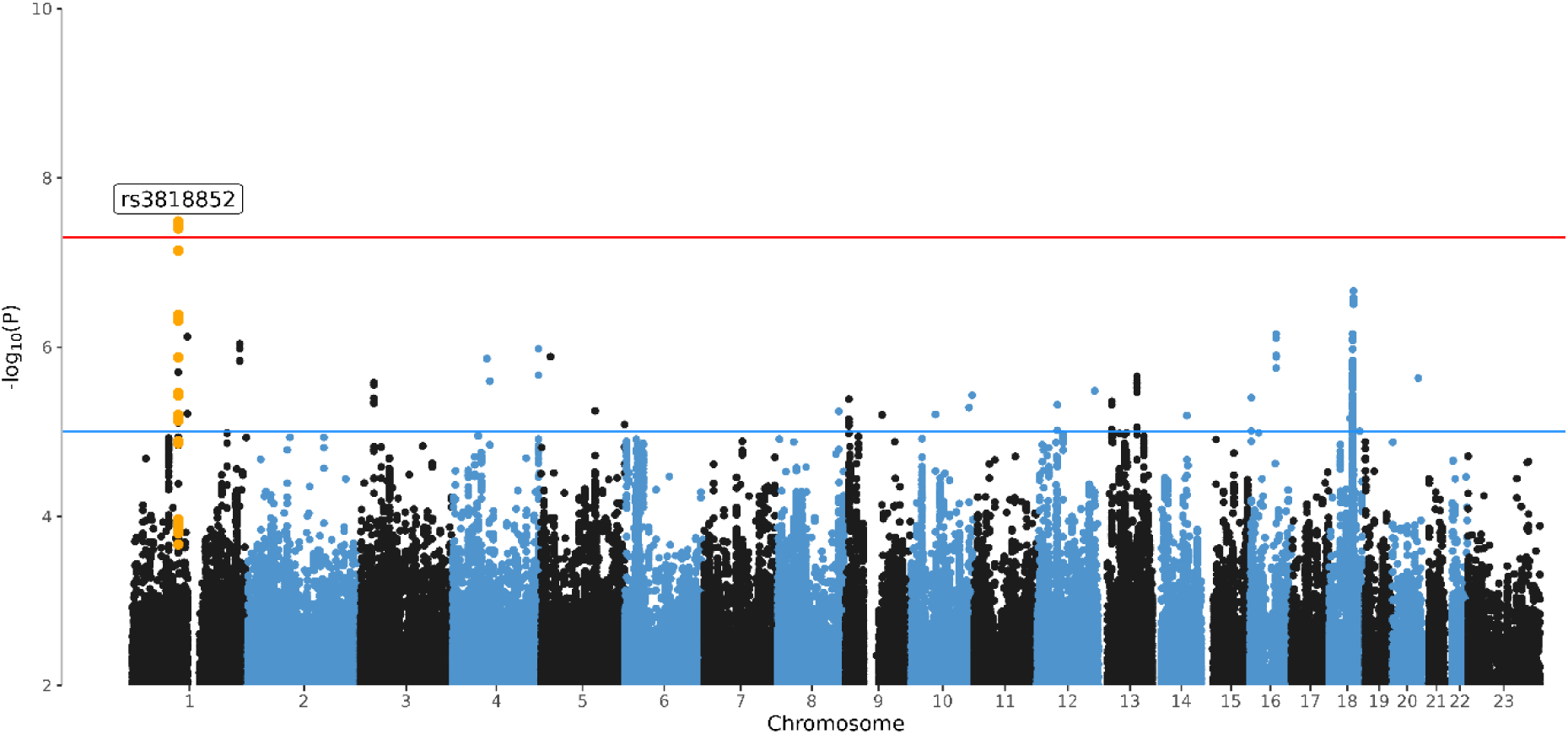
Manhattan plots of the GWAS mega-analysis of major depressive disorder. Manhattan Plot of the genome-wide association mega-analysis of lifetime MDD in BIONIC. The x-axis indicates chromosomal position. Y-axis denotes −log10 *p* and indicates the strength of association between the SNP and outcome phenotype. The red line indicates Bonferroni-corrected genome-wide significance; *p* < 5×10^-8^. The blue line indicates suggestive significance; *p* < 1×10^-5^.

### SNP-based heritability and genetic correlation with PGC-MD

We ran Linkage Disequilibrium score regression (LDSC)^30^ on the GWA mega-analysis results of lifetime MDD, hereafter called the BIONIC MDD GWAS, to compute SNP-based heritability (SNP-*h*^2^). SNP-*h²* for lifetime MDD in the effective sample size (N_eff_ = 63 743; N_cases_ = 16 655) was estimated to be 0.134 (se = 0.021) on the liability scale, suggesting 13.4% of phenotypic variance is jointly captured by the measured SNPs. This estimate is larger than those from recent large depression GWAS meta-analyses; e.g., the latest Psychiatric Genetics Consortium GWAS of major depression (PGC-MD) found SNP-*h*² = 8.4%, se = 0.07%^10^. This is consistent with empirical evidence that uniform phenotyping and clinical diagnostic criteria in a relatively homogeneous population benefit the capture of genetic signal of MDD^15,17^. We computed the genetic correlation between the BIONIC MDD GWAS and PGC-MD (European samples including 23andMe, N_eff_ = 1 523 738) in LDSC, finding a genetic correlation of *r*_G_ = 0.869 (se = 0.048).

### GWAS-by-subtraction

We investigated if a uniquely Dutch MDD signal might be identified through GWAS-by-subtraction^33^ (Figure 2A). The BIONIC MDD GWAS and PGC-MD GWAS were regressed on a latent factor representing genetic variance in depression aspects that are shared across the two studies, the residual being genetic variance that is unique to the BIONIC MDD GWAS, on which we ran a GWAS (Figure 2B). In this GWAS, no SNPs reached significance at the genome-wide adjusted alpha level. However, the rs3818852 SNP on chromosome 1 that was found in the BIONIC MDD GWAS remained among the top signals (OR = 0.83, *p* = 1.38 × 10^-6^). LDSC SNP-*h*^2^ for this model was 0.036 (se = 0.014).

**Figure 2.**
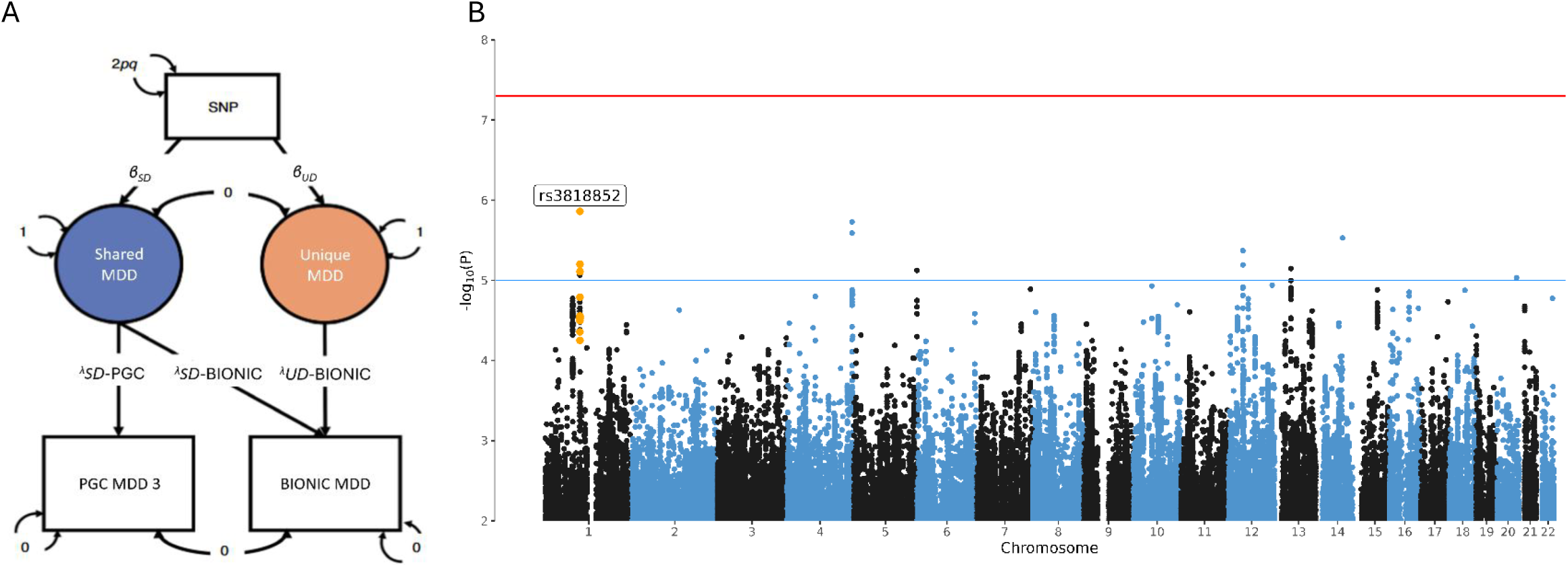
GWAS results of uniquely Dutch MDD (GWAS-by-subtraction). A) GWAS-by-subtraction model (Demange et al. (2021)). B) Manhattan Plot of the genome-wide association analysis of uniquely Dutch MDD, the latent factor resulting from subtracting the PGC-MD GWAS from the BIONIC MDD GWAS. The x-axis indicates chromosomal position. Y-axis denotes −log10 *p* and indicates the strength of association between the single-nucleotide polymorphism and the outcome phenotype. The red line indicates Bonferroni-corrected genome-wide significance; *p* < 5 × 10^-8^. The blue line indicates suggestive significance; *p* < 5 × 10^-5^.

### Polygenic score prediction

We tested predictive utility of the major depression polygenic score (MD PGS) based on PGC-MD in the BIONIC MDD GWAS sample (unrelated N = 44 144; 27% cases). The MD PGS was significantly associated with lifetime MDD status in BIONIC (OR = 1.054, *p* < 0.001), and explained 3.94% of variance on the liability scale. Figure 3A displays the increasing risk for MDD at higher MD PGS deciles.

**Figure 3.**
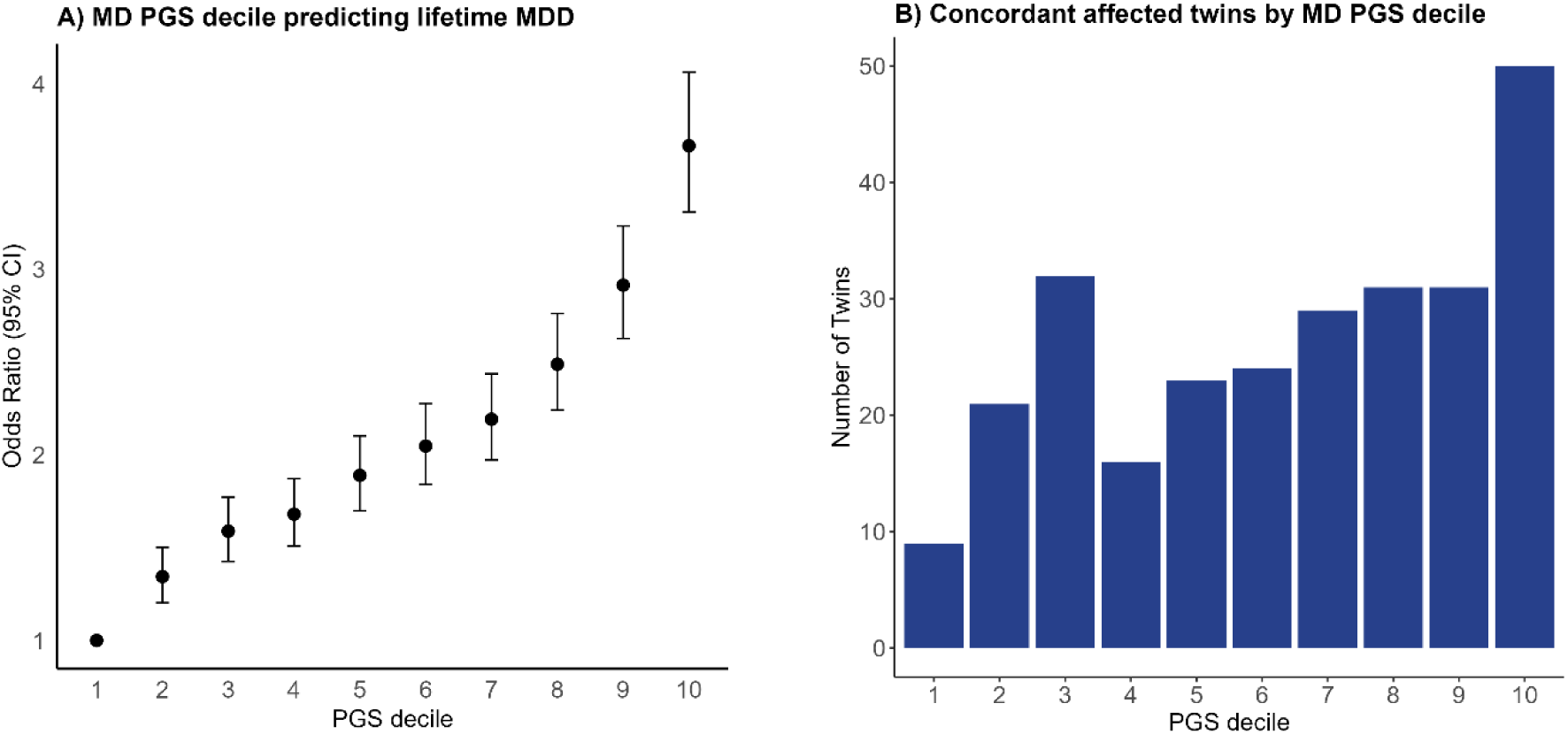
MD PGS prediction of lifetime MDD and twins by MDD PGS decile. A) Predictive PGS performance of MD PGS based on PGC-MD of lifetime MDD in BIONIC (N_unrelated_ = 44 144). The x-axis denotes the MD PGS decile. The y-axis denotes the odds ratio of each decile from the generalized estimating equations model where lifetime MDD status in the BIONIC sample was predicted by PGS decile. Odds ratios were estimated relative to decile 1. 95% confidence intervals are given. B) Number of twins concordant for lifetime MDD status per MD PGS decile (N = 266 twins).

We then tested predictive utility of a PGS based on the BIONIC MDD GWAS for two phenotypic definitions of depression in the UK Biobank: strict (ICD code for depression) and broad (‘minimal phenotypic’). The BIONIC MDD PGS was significantly associated with both strict (N = 23 755, OR = 1.163, Z = 17.1) and broad definitions of depression (N = 132 122, OR = 1.107, Z = 29.5), explaining 0.39% and 0.29% of phenotypic variance on the liability scale, respectively. These results highlight the transferability of the BIONIC MDD PGS, showing robust out-of-sample prediction for different phenotypic definitions.

#### Within- and between-family PGS prediction

Genotype-phenotype associations and the PGSs derived from them may arise through confounding effects, including population stratification, assortative mating, and gene-by-environment correlation^37^. To evaluate whether PGS prediction of lifetime MDD may be subject to confounding, we evaluated MD PGS performance based on PGC-MD in a subset of dizygotic twins from the BIONIC MDD GWAS sample. In an unmatched binary logistic regression model of 2282 dizygotic twins (1141 complete pairs), the major depression PGS significantly predicted lifetime MDD (OR = 1.434, *p* = 9.52 × 10^-9^). We then applied a random effects logistic model to predict lifetime MDD with between- and within-family MD PGS, and family as a random variable. Both the between- and within-family MD PGS were significantly associated with lifetime MDD (OR = 1.409, *p* = 9.04 × 10^-5^; OR = 1.948, *p* = 1.09 × 10^-6^, respectively). The between- and within-family PGS effects were significantly different (χ^2^ [1, N = 2282] = 3.983, *p* = 0.046), albeit barely so, suggesting that if there is confounding in MD PGS prediction, it is minimal. Sensitivity analyses showed that the confounding may in part be due to sex-specific effects, as the difference for the between- and within-family MD PGS effects was strongest in sex-discordant twins (*p* = 0.025), but absent in sex-concordant twin pairs (*p* = 0.435) (Supplementary Information).

#### Twin Concordance and PGS deciles

We computed MD PGS deciles in N = 2963 twin pairs in BIONIC. We distinguished three subsamples based on lifetime MDD concordance in the twin pairs: concordantly affected (N = 133 pairs), concordantly unaffected (N = 2257), and discordantly affected (N = 573). We expected concordantly affected twin pairs to be overrepresented in the high PGS deciles, and concordantly unaffected twin pairs to be overrepresented in the low PGS deciles. Figure 3B shows the distribution of concordantly affected twins across MD PGS deciles (concordantly unaffected and discordantly affected are shown in Supplementary Figure 4). Indeed, we found the number of concordantly affected twin pairs to increase in the higher MD PGS deciles, and the number of concordantly unaffected twin pairs to decrease in the higher MD PGS deciles. A similar pattern was observed in an independent replication sample from the Australian Genetics of Depression Study (Supplementary Information, Supplementary Figs. 5, 6).

We ran an ordinal logistic regression model with twin MDD concordance as outcome to test whether the mean MD PGS predicted twin MDD concordance status in the N = 2963 twins in BIONIC. We found each increase of one standard deviation in mean MD PGS to be significantly associated with a 1.3-fold greater risk for concordant MDD status within twin pairs (OR = 1.289; 95% CI = 1.174-1.404).

### Genetic causal associations

We computed genetic correlations between BIONIC lifetime MDD and 1461 complex traits and diseases, finding 388 significant genetic correlations (False Discovery Rate (FDR) < 5%) with well-established correlates including self-harm, substance use, anxiety, loneliness, neuroticism, irritable bowel syndrome, and chronic pain (Supplementary Material). With the latent causal variable (LCV) method^42,45^, we identified 33 traits with a putative causal genetic association with MDD at FDR < 5% (|genetic causal proportion (GCP)| > 0.60), and 4 traits with limited partial genetic causality (|GCP| ≤ 0.60) (Table 2; Figure 4). We identified 34 traits as putative risk factors for MDD and 3 traits as putative outcomes of MDD. The putative causal genetic associations with MDD include a diverse range of traits, including well-known risk factors such as hypersomnia, stroke, and trauma. Several significant putative causal traits were related to one’s work environment, including working with paints and chemicals and a workplace with very cold temperatures, which might also reflect differences in socio-economic status. Others included lifestyle traits such as vitamin D levels and vegetable consumption. Finally, associations with cardiometabolic traits were found, including stroke, ECG load, and HDL cholesterol.

**Figure 4.**
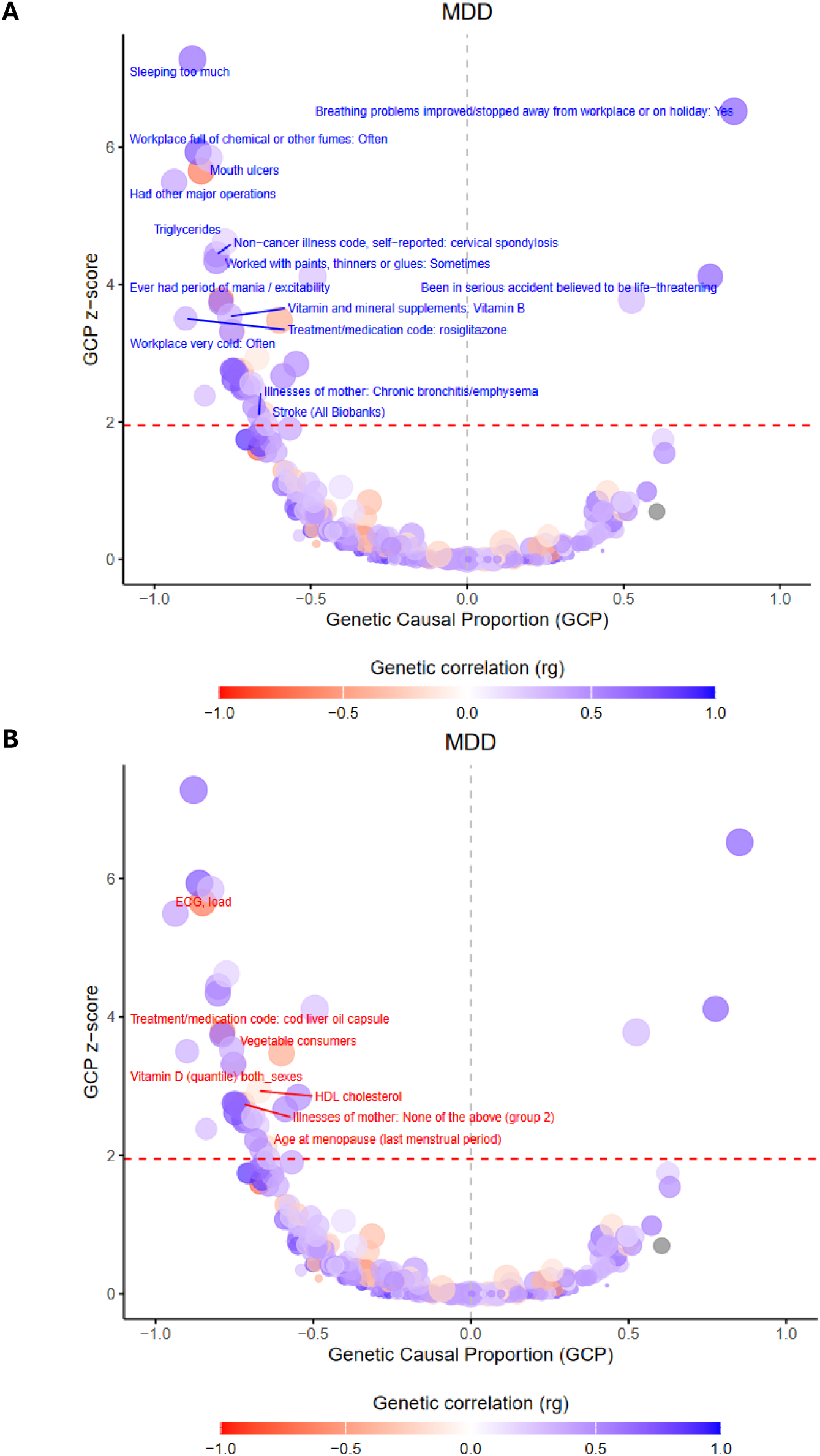
Causal architecture plots for MDD. Causal architecture plots depicting the LCV phenome-wide analysis results for BIONIC MDD. Dots represent traits with a significant genetic correlation with MDD. The x-axis reflects the proportion of genetic causality (GCP). The y-axis shows the GCP absolute Z-score (statistical significance). The red dashed line represents the statistical significance threshold (FDR < 5%), while the gray-dashed line represents the division between traits causally influencing MDD (left) and traits causally being influenced by MDD (right). Dot size reflects estimate accuracy (se). Annotation is provided for traits with a |GCP| > 0.60 that are significant at False Discovery Rate (FDR) < 5%, and that have a positive (plot A) or negative (plot B) genetic correlation with MDD.

### Gene-level analyses & tissue enrichment

We ran a MAGMA^43^ gene-based analysis on the BIONIC MDD GWAS results, mapping input SNPs to 19 210 protein-coding genes. Two genes were significantly associated with MDD at the genome-wide significance level: the *PALMD* gene on chromosome 1 (Z-score = 4.725, *p* = 1.15 × 10^-6^) and the *CIAPIN1* gene on chromosome 16 (Z-score = 4.627, *p* = 1.85 × 10^-6^) (Figure 5). We ran a MAGMA gene-set analysis for pathway-specific enrichment for gene sets C2 and C5 from MsigDB v7.0. We found no significant gene sets at the Bonferroni-corrected significance threshold (3.23 × 10^-6^). The most significant gene sets were involved in the regulation of T-cell migration (Beta = 1.61, *p* = 6.75 × 10^-6^) and vitamin binding (Beta = 0.31, *p* = 7.75 × 10^-6^). The full summary statistics of the gene-based test and gene set test are available in the Supplementary Material. In the MAGMA tissue expression analysis, in which we compared SNP associations from the BIONIC MDD GWAS with gene expression levels from the GTEx v8 database for 53 tissue types, none of the investigated tissues showed significant enrichment after multiple testing correction (Supplementary Figure 7).

**Figure 5.**
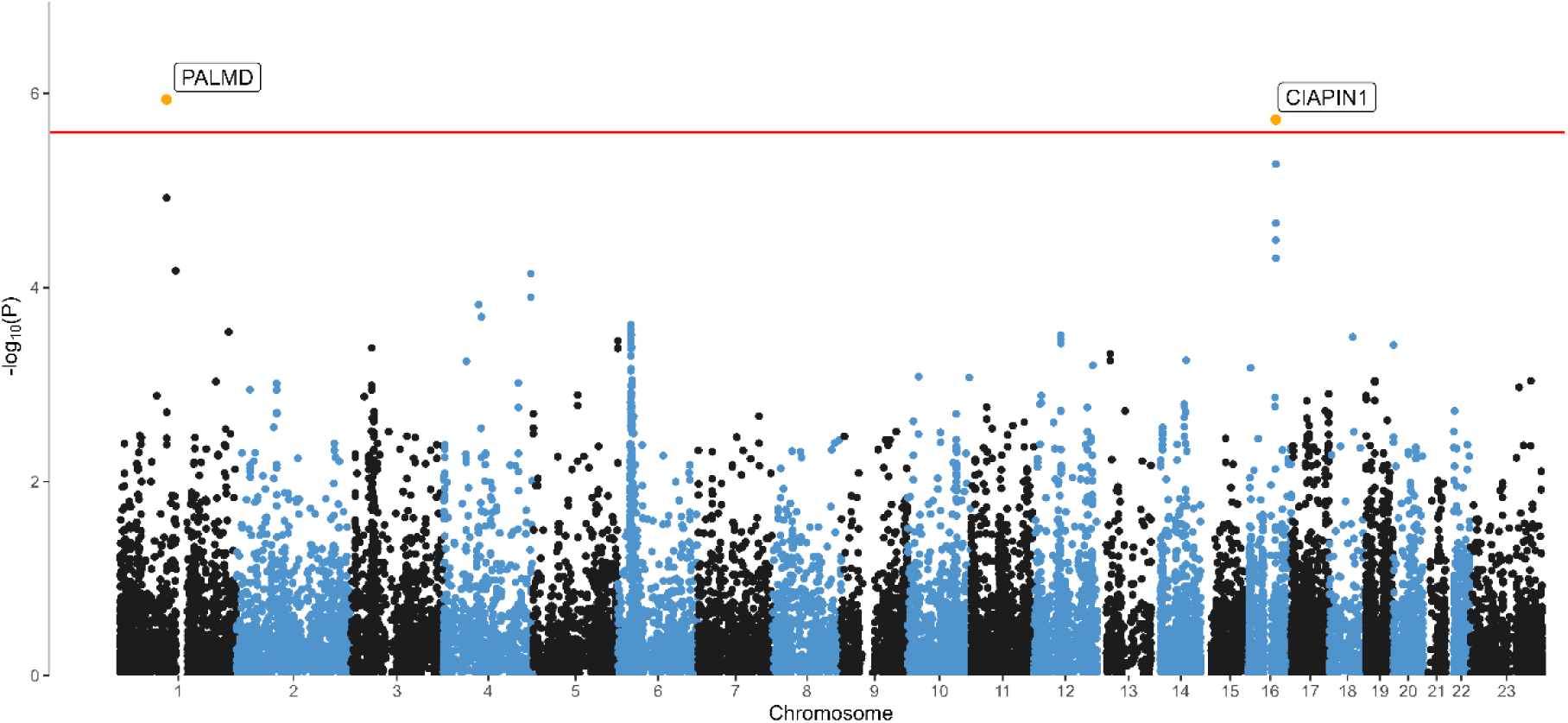
Gene-based analysis of MDD. Manhattan Plot of the gene-based analysis of lifetime MDD in BIONIC. The x-axis indicates chromosomal position. The y-axis denotes −log10 *P* and indicates the strength of association between the gene and the outcome phenotype. The red line indicates Bonferroni-corrected genome-wide significance; P < 2.50 × 10^-6^ (based on 19 994 tested protein-coding genes).

### Gene prioritization

We applied FLAMES^44^ to find the most likely effector gene for the genome-wide significant locus in the BIONIC MDD GWAS (lead variant: rs3818852 on chromosome 1). There were 23 SNPs with an LD > 0.60 implicated as potential alternative causal SNPs, which were used for credible set generation in FINEMAP. The *PALMD* gene was assigned the highest FLAMES score (0.436), suggesting it is the most likely effector gene for the genetic association between rs3818852 and MDD.

### Sensitivity analyses

To study the feasibility of a GWAS of MDD in a relatively small sample, we conducted a GWAS mega-analysis of height in 52 893 individuals from the BIONIC dataset. Despite the modest sample size, we identified 94 independent significant SNPs associated with height at the genome-wide significance level (Supplementary Figs. 8, 9). Height LDSC SNP-*h*^2^ was estimated to be 0.379 (se = 0.026; N_eff_ = 49 036), or 37.9% of phenotypic variance. The genetic correlation with the largest GWAS of height to date, Yengo et al.^46^, was *r*_G_ = 0.980 (se = 0.016).

As an alternative to the GWAS mega-analysis approach, we conducted a genome-wide association fixed effect meta-analysis combining the seven array group GWAS results of lifetime MDD case-control status. Results were similar to those of the main model (r_G_ = 1, Supplementary Information, Supplementary Figure 10).

**Table 1.** Traits with an inferred putative causal relationship with MDD.

| **Phenotypes that cause MDD** | **GCP** | **GCP se** | **GCP P** | **r<sub>G</sub>** | **r<sub>G</sub> se** | **r<sub>G</sub> P** |
| --- | --- | --- | --- | --- | --- | --- |
| Sleeping too much | -0.88 | 0.11 | 8.64e-16 | 0.65 | 0.16 | 4.01e-05 |
| Workplace often full of chemical or other fumes | -0.86 | 0.13 | 2.32e-11 | 0.73 | 0.19 | 7.56e-05 |
| Mouth ulcers | -0.83 | 0.13 | 5.31e-11 | 0.28 | 0.07 | 4.74e-05 |
| ECG load | -0.85 | 0.13 | 2.03e-10 | -0.68 | 0.18 | 1.68e-04 |
| Had other major operations | -0.94 | 0.15 | 6.11e-10 | 0.41 | 0.10 | 7.53e-05 |
| Triglycerides | -0.77 | 0.14 | 6.77e-08 | 0.20 | 0.07 | 6.87e-03 |
| Self-reported cervical spondylosis | -0.80 | 0.15 | 1.86e-07 | 0.44 | 0.15 | 2.38e-03 |
| Sometimes worked with paints, thinners or glues | -0.80 | 0.16 | 3.31e-07 | 0.54 | 0.12 | 1.18e-05 |
| Mouth-teeth dental problems: Mouth ulcers | -0.49 | 0.10 | 1.10e-06 | 0.28 | 0.08 | 2.49e-04 |
| Treatment/medication code: cod liver oil capsule | -0.79 | 0.17 | 4.97e-06 | -0.66 | 0.22 | 3.08e-03 |
| Ever had period of mania/excitability | -0.79 | 0.18 | 6.68e-06 | 0.60 | 0.12 | 5.69e-07 |
| Vitamin and mineral supplements: Vitamin B | -0.76 | 0.18 | 1.56e-05 | 0.36 | 0.13 | 6.57e-03 |
| Treatment/medication code: rosiglitazone | -0.90 | 0.21 | 1.89e-05 | 0.35 | 0.14 | 1.20e-02 |
| Vegetable consumption | -0.60 | 0.14 | 2.21e-05 | -0.43 | 0.17 | 1.30e-02 |
| Vitamin D | -0.75 | 0.18 | 4.35e-05 | -0.18 | 0.06 | 1.97e-03 |
| Workplace often very cold | -0.75 | 0.18 | 4.45e-05 | 0.51 | 0.15 | 4.61e-04 |
| HDL cholesterol | -0.67 | 0.18 | 1.73e-04 | -0.12 | 0.04 | 3.22e-03 |
| Workplace often very hot | -0.55 | 0.15 | 2.43e-04 | 0.50 | 0.15 | 5.84e-04 |
| Did your sleep change? | -0.75 | 0.21 | 3.44e-04 | 0.69 | 0.27 | 1.13e-02 |
| Difficulty concentrating during worst depression | -0.75 | 0.21 | 3.47e-04 | 0.75 | 0.14 | 1.60e-07 |
| Illnesses of mother: None of the above (group 2) | -0.72 | 0.20 | 3.75e-04 | -0.41 | 0.12 | 1.12e-03 |
| Workplace had a lot of cigarette smoke from other people smoking: Often | -0.74 | 0.21 | 4.57e-04 | 0.65 | 0.16 | 6.64e-05 |
| More creative or having more ideas than usual | -0.59 | 0.17 | 5.10e-04 | 0.51 | 0.17 | 2.24e-03 |
| Probable Recurrent major depression (moderate) | -0.75 | 0.22 | 6.30e-04 | 0.76 | 0.14 | 2.40e-08 |
| Diabetes diagnosed by doctor | -0.69 | 0.20 | 7.56e-04 | 0.24 | 0.07 | 3.16e-04 |
| Self-reported stroke | -0.72 | 0.22 | 8.71e-04 | 0.64 | 0.20 | 1.35e-03 |
| Treatment/medication code: ranitidine | -0.70 | 0.21 | 9.20e-04 | 0.46 | 0.14 | 8.28e-04 |
| Treatment/medication code: diazepam | -0.72 | 0.22 | 1.03e-03 | 0.63 | 0.24 | 1.00e-02 |
| Self-reported diabetes | -0.68 | 0.21 | 1.23e-03 | 0.23 | 0.07 | 1.38e-03 |
| Stroke (European biobanks) | -0.84 | 0.26 | 1.47e-03 | 0.30 | 0.11 | 8.61e-03 |
| Diaphragmatic hernia | -0.68 | 0.22 | 2.30e-03 | 0.49 | 0.14 | 3.52e-04 |
| Age at menopause (last menstrual period) | -0.65 | 0.22 | 2.99e-03 | -0.22 | 0.06 | 7.67e-04 |
| Illnesses of mother: Chronic bronchitis/emphysema | -0.67 | 0.23 | 3.42e-03 | 0.41 | 0.11 | 2.23e-04 |
| Stroke (global biobanks) | -0.64 | 0.23 | 4.74e-03 | 0.27 | 0.09 | 2.94e-03 |
| **Phenotypes caused by MDD** |  |  |  |  |  |  |
| Breathing problems improved/stopped away from workplace or on holiday | 0.85 | 0.12 | 3.55e-13 | 0.69 | 0.17 | 4.71e-05 |
| Been in a serious accident believed to be life-threatening | 0.78 | 0.16 | 1.08e-06 | 0.68 | 0.17 | 4.94e-05 |
| Back pain for 3+ months | 0.53 | 0.12 | 5.32e-06 | 0.31 | 0.11 | 6.39e-03 |

This table displays 37 traits with a significant (FDR < 5%) genetic causal proportion with lifetime major depressive disorder (MDD). P-values are before FDR correction; P-values after FDR correction are listed in Supplementary Material. GCP: genetic causal proportion; se: standard error; r<sub>G</sub>: genetic correlation.

Additionally, we conducted a GWAS mega-analysis of lifetime MDD using a less stringent screening process for psychopathology in control participants. This approach aimed to assess whether the screening criteria for controls introduced bias in SNP-h² and *r*_G_ estimates. Expanding the total sample size to N=67 440 (including 16 655 cases), we found that the relaxed control screening had minimal impact on the results (*r*_G_ = 0.99) compared to the main model; see Supplementary Information). The SNP-*h*² estimate in this model was 0.118 (se = 0.015), closely aligning with the estimate from the main model (SNP-*h*² = 0.134). These findings suggest that the SNP*-h*² estimate in the main model was not artificially inflated by the stringent control screening criteria.

## Discussion

We ran a genome-wide association (GWAS) mega-analysis of lifetime major depressive disorder (MDD) in the Dutch BIONIC dataset, including MDD and genotype data on 64 941 participants. We created this resource to help unravel the complex genetic architecture of MDD by focusing on uniform phenotyping based on DSM-5 criteria in a genetically homogenous sample.

We found that common SNPs explained 13.4% of variance in lifetime MDD on the liability scale. This is a notable estimate compared to recent large-scale meta-analyses using a combination of broad and diagnostic definitions (8.4%)^9–11^. Though the proportion of variance explained by common SNPs can be subject to variation due to many aspects of study design, a notable contributor is heterogeneity in population, study design, and phenotype. Tropf et al.^19^ found that heterogeneity across populations was one of the sources for the difference between twin-based heritability and SNP-based heritability, the so-called missing heritability, and Cai et al.^15^ found that restricting samples to those meeting clinical criteria benefitted explained genetic variance. Our findings corroborate empirical evidence that both sample uniformity and phenotypic precision can enhance genetic signal detection^17,19^, joining other recent efforts that focus on these aspects in gene discovery^47,48^. The success of our efforts reinforces the valuable contributions regional biobanks can have and suggests potential applications in studying other complex traits.

We report one genome-wide significant finding for MDD: rs3818852, an intronic variant of the *PALMD* gene on chromosome 1. To our knowledge, *PALMD* has not been implicated in MDD before. While its exact biological function is largely unknown, *PALMD* encodes the palmdelphin protein, which may be involved in cellular processes including cytoskeletal organization and cell shape modulation and is expressed in dendritic spines, membrane, and cytoplasm (NCBI, checked Feb 2025). Though rs3818852 has no previous associations, *PALMD* has been associated with complex traits from different phenotypic domains such as aortic valve calcification and the vaginal microbiome^49–51^. The A allele of the rs3818852 variant on chromosome 1 has a distinctly lower minor allele frequency in BIONIC (0.379) and in the Genome of the Netherlands reference panel (0.374)^22^ compared to the global population (0.486, TOPMED, https://www.ncbi.nlm.nih.gov/, checked Feb 2025). Differences in allele frequencies have been proposed as the leading reason for the success of the CONVERGE study in detecting two of the first GWAS loci for MDD^52^, neither of which were significantly associated with MDD in GWAS on European samples^9,11^. The *PALMD* signal was further reiterated by the GWAS-by-subtraction analysis, in which the signal remained after subtracting the global depression signal captured by the international PGC-MD effort. The gene-based analysis identified two significantly associated genes with MDD, corroborating the *PALMD* and finding the *CIAPIN1* gene. *CIAPIN1* is a protein-coding cytokine-induced inhibitor of apoptosis involved in mitochondrial function, methyltransferase activity, and iron-sulfur cluster binding. It has been found to inhibit hippocampal neuronal cell damage through MAPK and apoptotic signaling pathways^53^. To our knowledge, a role in MDD has not been established before.

Polygenic scores (PGSs) based on the BIONIC MDD GWAS significantly predicted distinct definitions of lifetime MDD in UK Biobank. The explained variance of the BIONIC GWAS PGS was higher for ICD10 diagnosis-based criteria, suggesting PGS based on strict phenotyping approaches may have higher transferability to other settings that use DSM criteria, such as the clinic. These results highlight that the BIONIC MDD PGS generalizes to other populations and phenotypic definitions. We also leveraged some of the unique features of the BIONIC dataset to explore polygenic score prediction of MDD in twin pairs. PGSs are increasingly becoming predictors and phenotypic proxies for phenotypes in genetic epidemiological designs^54^. GWAS and PGSs derived from these are naïve to how genotype-phenotype associations arise, meaning they can reflect biological mechanisms but also may capture environmental factors, and thus may be subject to confounding^37,55^. By leveraging features of twin- and within-family designs, we examined confounding in PGS prediction of MDD and found that if there was confounding due to population stratification, assortative mating, and gene-environment correlation, it was minimal, corroborating findings from previous work where estimates of assortative mating in MDD were low^20,56,57^. We also explored the association between twin concordance and polygenic burden following a recent example in schizophrenia and bipolar disorder^40^. We show that twin concordance for MDD is partly a function of polygenic burden, with concordantly affected twins having a higher PGS than pairs where one twin was affected, who in turn had a higher PGS than concordantly unaffected twins.

Estimates of genetic correlations and genetic causal associations with other traits identified several putative risk factors and outcomes of MDD. We found hypersomnia to increase the risk of MDD, together with workplace-related traits such as a cold workplace, working with fumes and paint thinners, having had major operations, and cardiovascular indicators like stroke and triglyceride levels. We found other traits to reduce the likelihood of MDD, including vitamin D levels, HDL cholesterol levels, vegetable consumption, and cod liver oil capsule consumption. MDD predicted that breathing problems improved when away from work, back pain lasting more than 3 months, and having been in a serious accident. In an earlier study of genetic causal associations with broad depression^45^, quite similar results were found, such as putative causal effects of hypersomnia, vitamin D levels, triglyceride levels, and various antidepressants. Conversely, the study found no putative outcomes of broad depression. It has been shown that minimally-defined depression includes nonspecific liability to psychopathology and that this affects genetic correlation estimates^15^. It is possible that this also affects genetic causal estimation estimates, and that considering clinically-defined MDD may reveal unique putative causal targets and outcomes.

There were several strengths and limitations. The sample size of this study was small compared to the standard of GWAS being published nowadays. Further, the results in this study were achieved by considering two factors, phenotypic homogeneity and population homogeneity. We cannot distinguish to which to attribute the increase in genetic signal, as indicated by the increase in SNP-*h*². Finally, it cannot go without mentioning that the lack of diversity in the sample might hinder the generalization of the findings to culturally-dependent phenotypic definitions and other ancestry-diverse populations. This study has been successful in recruiting cohorts that have not before been included in a GWAS of MDD, which we approached because they already had genotyping in the Dutch population available. We are not the first to show that a homogeneous and strict phenotyping definition benefits MDD genetic variant identification^52^. It is clear that such a strategy still holds promise for finding genome-wide significant associations with MDD in populations.

With this effort, we add to the growing evidence base on the genetic architecture of MDD. We show that an effort with uniform phenotyping in homogeneous populations can aid genetic signal detection for MDD, benefitting polygenic score prediction and the investigation of genetic correlates. As the genetic signal captured by broad phenotyping strategies plateaus, it may be time to invest in clinically relevant phenotyping approaches, for which we provide a proof of concept.

## Data Availability

Data are available upon reasonable request from the contributing cohorts. Full summary statistics from the BIONIC GWA mega-analysis of MDD can be found through the link below.

https://tweelingenregister.vu.nl/bionic-mdd-2025

