## Supplementary Information for "Genetics of Major Depressive Disorder in a Homogeneous Population with Uniform Phenotyping"

**BIONIC cohort descriptions**

**eMethods**

**Supplementary Figure 1**. Flowchart of the BIONIC GWAS MDD sample.

**Supplementary Table 1**. Demographic characteristics of the BIONIC GWAS sample (N = 64 941).

**Supplementary Figure 2.** QQplot of genome-wide association mega-analysis of lifetime MDD.

**Supplementary Figure 3.** Regional association plot of the PALMD region.

**Supplementary Table 2**. Top 10 independent SNPs in the genome-wide association mega-analysis of lifetime MDD. Positions and rsID are in build GRCh17.

**Within-family Polygenic Score Sensitivity models**

**Supplementary Figure 4.** Number of twins by MDD PGS decile for discordant affected and concordant unaffected pairs.

**Twin concordance and PGS quartile in the Australian Genetics of Depression Study and NTR**

**Supplementary Figure 5.** Number of twins by MDD PGS quartile and by twin pair MDD concordance in AGDS.

**Supplementary Figure 6.** Number of twins by MDD PGS quartile and by twin pair MDD concordance in NTR.

**Supplementary Figure 7.** MAGMA tissue enrichment analysis for MDD.

**Supplementary Figure 8.** Genome-wide association mega-analysis of height.

**Supplementary Figure 9.** QQplot of genome-wide association mega-analysis of height.

**Genome-Wide Association Meta-Analysis of MDD**

**Supplementary Figure 10.** GWAS meta-analysis of lifetime major depressive disorder case-control status.

**Additional screening of lifetime MDD controls**

**References**

### BIONIC cohort descriptions

*Doetinchem Cohort Study*

The Doetinchem Cohort Study (DCS) is a prospective population-based cohort study that started in 1987 and has been following over 12 000 participants in Doetinchem, the Netherlands, with follow-up of 8000 participants for over 30 years. The study aims to investigate the lifestyle and environmental determinants of chronic diseases and ageing in a life course perspective. Depression information was obtained using the LIDAS. For a detailed description of DCS, see Picavet, Blokstra, Spijkerman, & Verschuren^1^.

*The Hoorn Studies*

The Hoorn Studies comprise two population-based cohort studies (The Hoorn Study and The New Hoorn Study) initiated in 1989 and 2006 in Hoorn, the Netherlands. The studies aim to investigate the effects and interplay of genetic and environmental factors on a wide range of health outcomes, including cardiovascular disease, cancer, diabetes, and respiratory diseases. Participants have been followed prospectively and have been assessed at regular intervals on a range of clinical, lifestyle, and environmental factors. Biological samples have also been collected from participants at various time points. Depression information was obtained from the LIDAS. For a detailed description of the Hoorn Studies, see Rutters et al.^2^.

*The Hoorn Diabetes Care System cohort*

The Hoorn Diabetes Care System cohort is a prospective cohort study focused on studying the development and course of type 2 diabetes. Since its initiation in 1996 nearly 13 000 individuals have been included in the study with over 70 000 follow-up visits as of 2015. Participants with type 2 diabetes are followed over time with repeated measures on a range of demographic, physiological, lifestyle, clinical, mental health, and genetic outcomes. Depression information was obtained from the LIDAS. For a detailed description of the Hoorn Diabetes Care System cohort, see Heijden et al.^3^.

*Longitudinal Aging Study Amsterdam*

The Longitudinal Aging Study Amsterdam (LASA) is a prospective cohort study of older adults in the Netherlands. The study began in 1992 and includes over 5000 participants. Participants are assessed every 3 years on a range of physical, cognitive, emotional, and social functioning measures. The data also include genotype and mental health data. Depression information was obtained using the DIS across multiple measurement waves, the CIDI, and the CES-D. For a detailed description of LASA, see Hoogendijk et al.^4^.

*Lifelines*

Lifelines is a multi-disciplinary prospective population-based cohort study examining in a unique three-generation design the health and health-related behaviors of 167,729 persons living in the north of the Netherlands. It employs a broad range of investigative procedures in assessing the biomedical, socio-demographic, behavioral, physical and psychological factors which contribute to the health and disease of the general population, with a special focus on multi-morbidity and complex genetics*.* Depression information was obtained through the LIDAS instrument and the MINI v5.0. These data were either collected or prepared in van Loo, Aggen, & Kendler; van Loo et al.^5,6^. Lifelines genotype data were collected as part of Francioli et al.^7^. For a detailed description of Lifelines, see Sijtsma et al.^8^.

*MOod Treatment with Antidepressants or Running*

The MOod Treatment with Antidepressants or Running (MOTAR) study is a randomized controlled trial where depression and anxiety patients were assigned one of two treatments, antidepressant medication or running therapy, to investigate their impact on symptoms, biological aging and metabolic stress. Depression information was assessed using the CIDI across two measurement waves. For a detailed description of MOTAR, see Lever-van Milligen et al.^9^.

*MooDFOOD*

MooDFOOD is a 'Multi-country cOllaborative project on the rOle of Diet, Food-related behavior, and Obesity in the prevention of Depression'. It is a multidisciplinary consortium involving 13 organizations across 9 European countries, aimed at targeting food-related behaviors and nutritional status of people prone to overweight and obesity with the goal of preventing depression. The MoodFOOD sample is subclinical consisting of people with mild depressive symptomatology (PHQ-9 >= 5) but no current depressive episode, and BMI >= 25. For BIONIC, only the Dutch samples were included. Depression information was obtained at a baseline measurement with the MINI v5.0 and updated across three follow-up measurements, including one with where the LIDAS was administered. For a detailed description of the MooDFOOD project and its findings, see Bot et al.; Cabout et al.^10,11^.

*Nijmegen* Biomedische Studie

The Nijmegen Biomedische Studie (NBS) is a population-based study established in 2000 and expanded throughout several data collection waves. Set in the eastern part of the Netherlands, the study aims to investigate a wide range of demographic, clinical, biochemical, and genetic characteristics in the general Dutch population. Depression information was obtained through the LIDAS instrument. For a detailed description of NBS, see (Galesloot et al.^12^.

*Netherlands Study of Depression and Anxiety*

The Netherlands Study of Depression and Anxiety (NESDA) is a longitudinal cohort study that aims to investigate the etiology, course, and consequences of depressive and anxiety disorders. The study began in 2004 and includes over 3300 participants with a current or remitted depressive or anxiety disorder, as well as their siblings and healthy controls. Participants are assessed every 2 years on a range of clinical, psychosocial, and biological measures. Depression information was obtained through the CIDI across several measurements. For a detailed project description, see Penninx et al.^13^.

*Netherlands Study of Depression and Anxiety sibling cohort*

The Netherlands Study of Depression and Anxiety sibling cohort (NESDAsib) is a side branch of the NESDA project comprising 367 full siblings of the NESDA cohort. Depression information was obtained through the CIDI. For a detailed description of NESDAsib, see Penninx et al.^13^.

*Netherlands Study of Depression in Older Persons*

The Netherlands Study of Depression in Older Persons (NESDO) is a prospective cohort study that aims to investigate the determinants, course, and consequences of depression in older persons. The study began in 2007 and includes over 500 participants aged 60 years and older with a current or recent diagnosis of depression. Participants are assessed every 2 years on a range of clinical, psychosocial, and biological measures. The study has been used to study the impact of depression on physical health, cognitive function, and quality of life. NESDO findings have been used to develop and evaluate interventions to improve the lives of older adults with depression. Depression information was obtained using the CIDI across three measurement waves. For a detailed description of NESDO, see Comijs et al.^14^.

*Nutrition Questionnaires plus*

The Nutrition Questionnaires plus (NQplus) study is a prospective cohort study that aims to investigate the dietary determinants of cardiometabolic health in Dutch adults. The study began in 2011 and includes over 2000 participants aged 20–70 years. Participants are assessed every 2 years on a range of dietary and health measures, including mental health. Depression information was obtained using the LIDAS. For a detailed description of NQplus, see Brouwer-Brolsma et al.^15^.

*Netherlands Twin Register*

The Netherlands Twin Register (NTR) is a population-based cohort of over 200 000 twins and twin-families from across the Netherlands. NTR respondents are periodically invited to participate in lab and questionnaire measurements, resulting in a vast array of biological and behavioral data. NTR-Biobank^16,17^ includes around 10 000 participants with genotyping and biomaterials. Additional genotype either in blood or buccal DNA is available in an extra 15 000 samples. Depression information was obtained from the LIDAS, the CIDI, and the ASR-ASEBA, BDI, and HADS. For a detailed description of the NTR, see Ligthart et al.^18^.

*Tracking Adolescents’ Individual Lives Survey*

The Tracking Adolescents’ Individual Lives Survey (TRAILS) and TRAILS clinical cohort (TRAILS-CC) are a prospective population-based and clinical cohort study of adolescents in the Netherlands. The studies aims to investigate the development of adolescents into young adults, with a focus on mental health, social-emotional development, and health behaviors. The population cohort consists of N = 2230 and TRAILS-CC of N = 543 participants who were born in 1996 or 1997; participants have been followed prospectively since they were 11 years old. Depression information was obtained using the LIDAS. For a detailed description of the TRAILS and TRAILS-CC studies, see Oldenhinkel et al.^19^.

### eMethods

#### The BIONIC study

The BIONIC (BIObanks Netherlands Internet Collaboration) project includes sixteen cohort studies from across the Netherlands united in establishing a depression phenotype and genotype database to study the etiology and characteristics of depression in the Dutch population^20^. When an major depressive disorder (MDD) definition based on the Diagnostic and Statistical Manual of Mental Disorders (DSM-5)^21^ was not already available, uniform data collection was realized through an online diagnostic instrument, the Lifetime Depression Assessment Survey (LIDAS)^22^, developed to assess DSM-5 depression. These efforts resulted in a dataset of 132 402 Dutch individuals with information on lifetime MDD diagnosis, depression symptom status, episode characteristics, and a range of clinical and demographic indicators, including educational attainment, smoking, body-mass index and medication use. At the time of this study, 72 315 of these participants also had genome-wide single nucleotide polymorphism (SNP) data; of this group 64 941 had MDD case-control status, all covariates, good quality genotype data and were of European ancestry (Supplementary Figure 1).

#### Genotype data

The sixteen cohorts had collected genotype data with different arrays, resulting in 19 datasets where the same arrays could be available in different cohorts, or where the same cohort had performed genotyping on different arrays. For quality control (QC), samples were excluded in case of discrepancy between reported and biological sex (PLINK F_chrX_-coefficient < 0.8 for males and F_chrX_ > 0.2 for females), excess heterozygosity (F_autosomes_ > 0.10 or < -0.10), insufficient sample call rate (< 0.90) and call rate by chromosome (< 0.80), or incorrect identity-by-descent sharing between relatives. SNPs were excluded based on Hardy Weinberg Equilibrium (p < 1x10-4) and SNP call rate (< 0.95), as well as Mendelian error rates above 1%. Palindromic SNPs were excluded with minor allele frequency (MAF) > 0.30. SNPs were aligned to the Haplotype Reference Consortium (HRC) reference panel (v1.1)^23^ and SNPs with an allele frequency difference > 0.10 with the reference data were excluded. Some of the datasets did not fit independent GWAS analysis criteria because they were too small or had an overrepresentation of cases. To include the largest number of individuals, SNP data genotyped on the same arrays were combined. This resulted in seven array groups with raw genotype data from Affymetrix 6, Axiom Finngen, Axiom-NL, Illumina CytoSNP, Global Screening Array, Human Core Exome, and Omnichip. These array groups were the units for further QC, including duplicate sample removal, a repeat of the SNP QC thresholds above, and a MAF threshold of 0.01. Genotype data were then imputed against the HRC reference panel (v1.1). Further details on the quality control procedure are provided elsewhere^20^. After imputation, the seven array groups had identical genome coverage and were merged into a single set for mega-analysis. All arrays included genotype data on the 22 autosomes. The availability of X-chromosome data varied (Supplementary Figure 1), with the first pseudo-autosomal region (PAR), non-PAR, and second PAR available in 55 141, 55 863, and 41 441 individuals, respectively.

We conducted principal component analysis (PCA) in PLINK (v1.9)^24^ on the imputed data based on the three superpopulations (African, Asian, European) from the 1000 Genomes Project reference panel (phase 3v5)^25,26^. The BIONIC genotype data were SNP and LD (linkage disequilibrium) pruned and projected onto the 1000 Genomes Project PCA space to compute principal components (PCs). Ancestry outliers in the BIONIC data were excluded based on a 4 SD distance from the mean of 0 for the first 6 standardized PCs. Additional ancestry outliers were removed based on visual inspection of PC clustering.

#### Phenotype data

Lifetime MDD cases were uniformly defined according to DSM-5 diagnostic criteria as ever having had a period of at least two weeks with five or more depression symptoms causing dysfunction in life, of which at least one a cardinal symptom^21^. Lifetime MDD cases were primarily (47%) identified through the Lifetime Depression Assessment Survey (LIDAS)^22^. This online self-report questionnaire was developed to reliably and efficiently assess MDD diagnosis for large population studies and ongoing biobank studies based on the Diagnostic and Statistical Manual 5 diagnostic criteria^21^. The LIDAS was validated against the composite international diagnostic interview (CIDI)^27^ and has been used in a number of epidemiological and genetic studies since^5,28–31^. The remaining diagnoses were derived from the CIDI, mini international neuropsychiatric interview (v5; MINI)^32^, and diagnostic interview schedule (DIS)^33^.

The majority (87%) of lifetime MDD controls were defined in the absence of 5 symptoms, a core symptom, or dysfunction. Additional lifetime MDD controls (13%) were identified based on low scores on one or multiple depression symptom instruments, including the BDI, CES-D, HADS, and ASR-ASEBA^34^. As an additional screener, lifetime MDD controls were asked about diagnostic and treatment history for a range of psychopathologies when such information was available (56%). This included screening for a self-reported diagnostic or treatment history of depression, anxiety disorder, bipolar disorder, any eating disorder, schizophrenia, obsessive compulsive disorder, post-traumatic stress disorder, phobia, attention-deficit or hyperactivity disorder or attention deficit disorder, any personality disorder, alcohol or drug addiction, and use of antidepressant medication.

Age was defined as years at MDD assessment. If multiple measures for the same person were available, the timepoint was chosen where diagnostic criteria for MDD were first met (for cases), or the latest measurement at which diagnostic criteria had not been met (for controls). Individuals with missing age information or ages outside the range of 16 to 105 were excluded. Sex was defined based on self-report and confirmed through genotyping.

#### Polygenic scores

##### Predicting lifetime MDD status in BIONIC with polygenic scores

We generated major depression polygenic scores (PGSs) in BIONIC based on the PGC-MD GWAS summary statistics (European samples), excluding all datasets from the Netherlands^35^. PGSs reflect the genetic propensity for a trait based on the weighted sum of risk increasing and decreasing alleles across the genome, with allelic weights derived from GWAS. PGSs were generated in the LDpred software (v0.9.1)^36^, a Bayesian method that derives posterior mean causal effect sizes from GWAS summary statistics by assuming a prior for the genetic architecture and linkage disequilibrium (LD) information. For the prior, an infinitesimal model was assumed. LD block information for West-European ancestry was derived from the UK Biobank^37^. The association between the MD PGS and lifetime MDD case-control status in BIONIC was estimated by logistic regression in unrelated individuals with age at assessment, sex, and 10 ancestry informative PCs as covariates. The proportion of variance explained by the PGS on the liability scale for MDD case-control status was estimated according to Lee et al.^38^.

##### Out-of-sample prediction

We explored out-of-sample MDD PGS prediction based on the BIONIC MDD GWAS summary statistics in the UK Biobank. Genetic and clinical data from the UK Biobank were accessed under application number 16406^37^. The UK Biobank initiative has been approved by the National Research Ethics Service Committee (reference 11/NW/0382). The imputation and quality control performed by the UK Biobank team together with additional in-house genomic control are described extensively elsewhere^37,39^. After QC, a total of 9 380 668 high-quality SNPs were hard-called under a certainty threshold of 0.9^40^. Analyses were performed on common (MAF > 0.01) genotyped SNPs or common SNPs with very high imputation quality (INFO>0.9). Principal components from the 1000 Genomes reference populations^41^ were projected onto the called genotypes available using FLASHPCA2^42^. Subjects with inferred European ancestry were kept for downstream analyses. Ancestry outliers (6 standard deviations to the average score of Europeans) were excluded. Subjects with sex aneuploidy, discordant reported and chromosomal sex, and relatives were excluded from the analysis. The final sample size resulted in 387 553 unrelated individuals of European ancestry.

Two depression phenotypes were defined based on the clinical data available for the UK Biobank cohort. First, a strict phenotype definition was constructed based on diagnosis of single or recursive depressive episodes (ICD10 codes: F32, F33) present in the hospital records. Next, a broad definition of depression based on Howard et al.^43^ was adopted. A case was defined by answering yes to one of two questions in at least 1 assessment time point: ‘Have you ever seen a general practitioner for nerves, anxiety, tension or depression?’ (data field: 2090) or ‘Have you ever seen a psychiatrist for nerves, anxiety, tension or depression?’ (data field: 2100) or meets criteria for strict definition. Subjects diagnosed with schizophrenia, schizoaffective or delusional disorders (F20-F29) or bipolar disorder (F31) were excluded from the analysis. A total of 23 755 and 132 122 individuals met the criteria for strict and broad phenotype definitions, respectively.

PGSs were calculated using BIONIC MDD GWAS summary statistics similarly to above. PGSs were subset for alleles present in both datasets and reference alleles were aligned. HAPMAP3 SNPs were kept for downstream analyses. Scores were calculated using LDpred (v1.0.10)^36^. First, data were synchronized using a subsample of 10 000 unrelated European individuals from the UK Biobank. Then, the SNP weights were calculated for the target sample using a LD radius of 100. The weights from the infinitesimal model were then used to perform allele scoring on the target sample with PLINK (v1.9). To examine the predictive power of the BIONIC MDD PGS in the UK Biobank, a logistic regression model was fitted. The primary independent variable was the standardized PGS. Sex, age, genotyping array, and the first 10 principal components were added as covariates.

##### Within-family prediction

PGS prediction of a trait can be subject to confounding, including bias from population stratification and assortative mating, as well as passive gene-environment correlation, where genes do not only directly affect our phenotype but also our (rearing) environment through the genotypes of our parents^44–46^. Within-family designs are able to account for many sources of passive genotype-environment correlation, with dizygotic (DZ) twins having the additional benefit that all shared environmental factors are time-invariant among twins^47^.

We sought to evaluate MD PGS prediction in a within-family design, following the approach of Selzam et al.^47^ with DZ twins from the Netherlands Twin Register (NTR; a collaborating cohort in BIONIC). We identified 1141 dizygotic twin pairs with non-missing lifetime MDD case-control status and MDD polygenic scores. First, PGS prediction of MDD was assessed in the entire DZ twin sample through a logistic generalized linear mixed-effects model, correcting for sex and genotype array. Next, between-family PGS effects were defined as the change in lifetime MDD risk with a change in average family PGS values, and within-family PGS effects as the change in lifetime MDD risk with a change in the difference between individual PGS and the family average PGS. The two were fit as predictors of MDD in the same model, together with a random effect for family, so that individual estimates are adjusted for and independent of the effect of the other estimate. The statistical difference between the between- and within-family PGS effects was then empirically tested through a chi-square test of the difference between their coefficients divided by the standard deviations of the sampling distribution of the estimate differences^48,49^. A statistical difference here would indicate that the PGS association with a trait may be partly explained by confounding. We conducted sensitivity analyses where data were restricted to sex-concordant DZ twin pairs (N = 1408; 704 pairs), sex-discordant DZ twin pairs (N = 874; 437 pairs), and DZ twin pairs in which twins were genotyped on the same array (N = 2084; 1042 pairs).

##### Polygenic scores and twin concordance

We computed MD PGS deciles based on the PGC-MD summary statistics in N = 2963 complete twin pairs from the NTR cohort who took part in BIONIC. We then distinguished 3 groups: concordantly affected (N = 133 pairs), concordantly unaffected (N = 2257), and discordantly affected (N = 573). We hypothesized that concordantly affected twins would be overrepresented in high MDD PGS deciles. We repeated this approach in an independent sample for replication, the Australian Genetics of Depression Study^50^. We formally tested the association between the mean MD PGS in NTR twin pairs and MDD concordance in an ordinal logistic regression model, correcting for two ancestry-informative PCs^51^.

#### Genetic correlations

We computed genetic correlations between the BIONIC lifetime MDD GWA mega-analysis and 1461 disease, personality and lifestyle traits using bivariate LDSC in the Complex-Traits Genetics Virtual Lab (CTG-VL) analysis pipeline (<https://vl.genoma.io/>). CTG-VL is a virtual web-tool that houses a collection of 1461 GWAS summary statistics for free public access and post-GWAS analyses^52^. The GWAS results were derived from GWAS consortia and the second wave of GWAS results by the Neale Lab ([www.nealelab.is/uk-biobank/](http://www.nealelab.is/uk-biobank/)). The CTG-VL platform incorporates GWAS results predominantly from European ancestry samples. The inclusion criteria for GWAS in CTG-VL includes having a nominally significant LDSC SNP-heritability (P-value < 0.05 before multiple testing correction). These GWAS were adjusted for age, age-squared, inferred chromosomal sex, age * inferred sex, age-squared * inferred sex, and 20 genetic ancestry-informative principal components^52^.

#### Latent Causal Variable analysis

We applied the bivariate latent causal variable (LCV) model to the 388 traits from the CTG-VL pipeline that had a significant genetic correlation with the BIONIC MDD GWAS. The LCV method is a statistical approach to identify potentially causal relationships among heritable traits^53^. Specifically, it quantifies the degree to which a genetic correlation between two traits may be explained by (partial) genetic causality as opposed to full horizontal pleiotropy. In the LCV model, a genetic correlation between trait A and B is mediated by a latent variable (L) that represents the causal component between the two traits. The core metric used in this method is the genetic causal proportion (GCP), which is derived from correlations between trait A and L, and trait B and L, and quantifies the proportion to which a genetic correlation can be explained by potential causal effects. Assuming no bi-directional causality, a GCP value of 0 suggests the genetic correlation is purely due to horizontal pleiotropic effects, whereas a |GCP| of 1 indicates complete genetic causality. Intermediate GCP values denote partial genetic causality, where some, but not all, of the genetic correlation can be explained by causal effects. A |GCP| > 0.60 is considered to be robust^54–56^. A negative GCP would suggest that a given trait is likely to be a risk or protective factor for MDD, whereas a positive GCP would suggest that MDD causes the trait. Benjamini-Hochberg’s False Discovery Rate < 5% was applied to correct for multiple testing at both the genetic correlation and LCV steps.

#### Gene analysis & tissue enrichment

We performed a gene-based test and gene-set analysis of MDD in the MAGMA (Multi-marker Analysis of GenoMic Annotation; v1.08)^57^ tool. MAGMA employs a multiple linear principal components regression and F test to obtain P values for genes^57^. In the gene-based test, input SNPs from the BIONIC MDD GWAS were mapped to 19 210 protein coding genes (gene window 10kb), and tested for association with MDD in a SNP-wise top model. Analyses were adjusted for possible confounders such as gene size, gene density, linkage disequilibrium and minor allele count. Genome-wide significance was set at the Bonferroni-corrected threshold of P = 0.05 / 19 210 = 2.6 × 10^-6^. Pathway-specific enrichment of genetic signal for MDD was explored in competitive gene-set analysis with the gene-based analysis of MDD as input. Association between gene-set membership and gene Z-scores was computed using MAGMA. Gene sets were derived from categories C2 and C5 from the publicly-accessible MsigDB v7.0. Significance was set at the Bonferroni-corrected threshold of P = 0.05 / 15 488 = 3.23 × 10^-6^. We performed tissue expression analysis in MAGMA with the gene-based test results as input and gene expression datasets obtained from GTEx v8 RNAseq. The MAGMA gene-property test compares average gene-expression for 53 tissue categories conditioning on average expression across all categories (one-side) to test for tissue-specific enrichment. Significance was defined at P = 0.05 / 18 062 = 2.77 × 10^-6^.

#### FLAMES gene prioritization

We fine-mapped the BIONIC MDD GWAS results in the Fine-mapped Locus Assessment Model of Effector geneS (FLAMES)^58^ software. FLAMES integrates machine learning predictions linking SNPs to genes with GWAS-wide convergence of gene interactions, to predict the most likely effector gene (i.e., the gene through which a SNP enacts its effect on the phenotype) in a locus. Significant loci in the BIONIC MDD GWAS were subjected to FLAMES to determine the effector genes. Gene Z-scores and tissue enrichment estimates were derived from the MAGMA gene-based test and tissue expression analyses, respectively, and served as input for the Polygenic Priority Score (PoPS)(v0.2)^59^ tool. An LD matrix was generated based on the BIONIC MDD GWAS sample in LDstore2 (v2)^60^ and posterior probabilities for 95% credible sets of causal SNPs were derived from the MDD GWAS summary statistics using FINEMAP (v1.4)^61^. Information from the MAGMA, PoPS and FINEMAP analyses were combined in FLAMES to generate the FLAMES score, an estimate between 0 and 1 assigned to genes near the associated locus where the highest estimate indicates the most likely effector gene.

#### Sensitivity analyses

Height, having a well-characterized genetic architecture, was used a benchmark trait to compare the results form lifetime MDD to. To this end, we conducted a GWAS mega-analysis of height in N = 52 893 Dutch individuals from the BIONIC study. Height in centimeters was obtained via self-report. Outlying values (≤ 140 and ≥ 210) were excluded. Analyses were conducted in fastGWA^62^ from the Genome-wide Complex Trait Analysis (GCTA) software^63^, with mixed linear model. SNPs with a minor allele frequency below 0.01 and imputation quality below 0.40 were excluded from analysis. Analyses were corrected for sex, age, 10 ancestry-informative principal components, genotype array and relatedness (through a genetic relationship matrix). SNP-*h*^2^ and *r*_G_ estimates were calculated in LDSC^64^.

We conducted a fixed-effect genome-wide association meta-analysis of lifetime MDD in N = 64 941 Dutch individuals (16 655 cases) from the BIONIC study. First, genome-wide association analyses of lifetime MDD were conducted in in the seven array groups that make up the BIONIC genotype data (specified in eMethods). GWAS were conducted in fastGWA^62^ from the Genome-wide Complex Trait Analysis (GCTA) software^63^, using a generalized linear mixed model. Except for genotype array, the same covariates as in the main analysis were applied: sex, age, and 10 ancestry-informative principal components. We computed a genetic relatedness matrix (GRM) in GCTA to address any relatedness in the sample. For computational efficiency, GRM values below 0.05 were set to 0 in a sparse GRM, which was included in the association analyses. SNPs with an allele frequency below 0.01 were excluded from analysis. GWAS results were filtered on imputation quality ≥ 0.40 and Hardy Weinberg Equilibrium test p-value ≤ 0.0001 and were meta-analyzed with the Genome-Wide Association Meta-Analysis (GWAMA) software^65^. SNP-*h*^2^ and *r*_G_ estimates were calculated in LDSC^64^.

In the main GWAS model, lifetime MDD controls (as defined through DSM-5 criteria or low scores on depression questionnaires) were screened for a history of diagnosis or treatment for a range of psychopathology and antidepressant use. Lifetime MDD controls who reported ‘yes’ to any of these measures were excluded from analyses. Although common for MDD, screening controls for any psychiatric disorder can lead to the selection of super-controls, i.e., individuals who are healthier than expected by chance^66^. Such controls could be especially depleted for deleterious variants, violating the random effects assumption of GWAS. This would, in turn, lead to biased SNP-based heritability (SNP-*h*²) and genetic correlation (*r*_G_) estimates. Here we considered the effect of additional screening of lifetime MDD controls on SNP-*h*² and *r*_G_ with the main model and PGC-MD results. For this we defined an alternative phenotype definition where lifetime MDD controls were screened only for depression and not for additional psychopathology. We ran a GWAS mega-analysis of MDD in 67 440 individuals (16 655 lifetime MDD cases and 50 785 controls) in fastGWA^63^ using identical parameters and covariates as the main model (sex, age, genotype array, 10 ancestry-informative PCs, and relatedness). SNP-*h*^2^ and *r*_G_ estimates were calculated in LDSC^64^.

### **Supplementary Figure 1**. Flowchart of the BIONIC GWAS MDD sample.

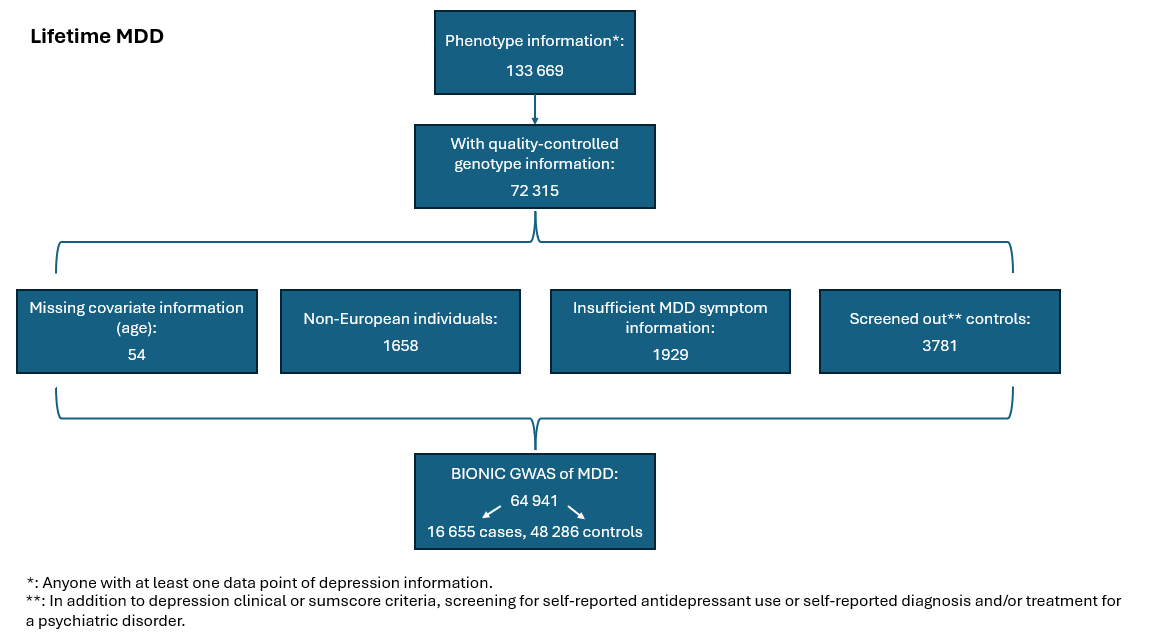

### **Supplementary Table 1.** Demographic characteristics of the BIONIC MDD GWAS sample (N = 64 941).

|  | **Full sample**  **N = 64 941** | **MDD cases**  **N = 16 655** | **MDD controls**  **N = 48 286** |
| --- | --- | --- | --- |
| Age (mean years, SD) | 50.7 (16.0) | 48.7 (14.3) | 51.4 (16.5) |
| Sex, % female | 60.8% | 70.4% | 57.4% |

MDD = major depressive disorder.

### **Supplementary Figure 2.** QQplot of genome-wide association mega-analysis of lifetime MDD.

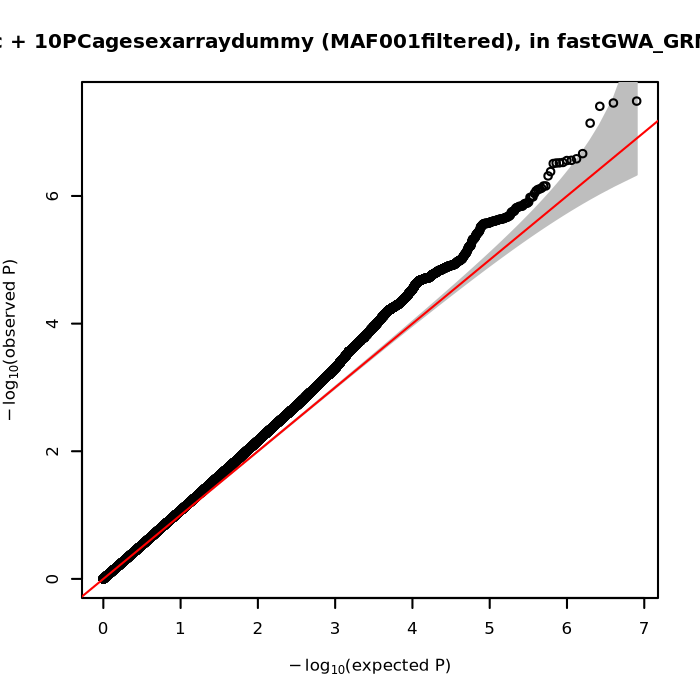

QQplot showing expected versus observed -log10 P-values from the genome-wide association mega-analysis of lifetime major depressive disorder (MDD) (N = 64 941).

### **Supplementary Figure 3.** Regional association plot of the PALMD region.

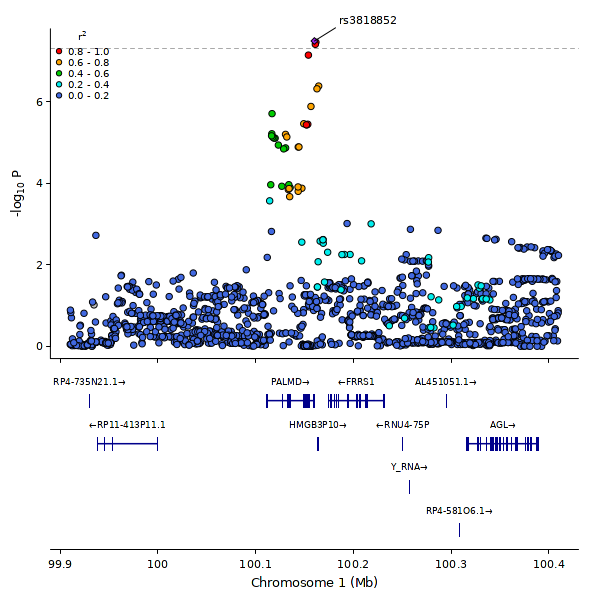

Regional association plot of the PALMD region, identified in the GWAS mega-analysis of lifetime MDD. The y-axis shows the -log10 p-values of the SNPs. The x-axis represents their chromosomal positions. Arrows mark the genomic locations of genes within the regions of interest based on the NCBI Build 37 human assembly. SNP colors indicate linkage disequilibrium with the top associated SNP. The figure was generated using LocusZoom (<http://csg.sph.umich.edu/locuszoom/>).

### **Supplementary Table 2**. Top 10 independent SNPs in the genome-wide association mega-analysis of lifetime MDD. Positions and rsID are in build GRCh17.

| **Chr** | **SNP** | **BP** | **A1/A2** | **F(A1)** | **OR** | **s.e.** | ***P*** | **Nearest Gene** |
| --- | --- | --- | --- | --- | --- | --- | --- | --- |
| **1** | **rs3818852** | **100160177** | **A/T** | **0.38** | **0.93** | **0.01** | **3.26e-8** | ***PALMD*** |
| 18 | rs4131791 | 52747871 | C/T | 0.58 | 1.07 | 0.02 | 2.17e-7 | LINC3035 |
| 18 | rs12327102 | 50787839 | G/C | 0.46 | 0.93 | 0.01 | 6.96e-6 | DCC |
| 16 | rs62037142 | 57477652 | C/T | 0.98 | 0.76 | 0.04 | 6.97e-6 | *CIAPIN1* |
| 1 | rs150207136 | 119607219 | G/A | 0.98 | 0.79 | 0.04 | 7.53e-6 | WARS2 |
| 1 | rs1970168 | 232761116 | G/A | 0.51 | 1.07 | 0.02 | 9.13e-6 | SIPA1L2 |
| 4 | rs10000918 | 186268576 | A/G | 0.74 | 1.08 | 0.02 | 1.04e-6 | SNX25 |
| 5 | rs33935044 | 21145325 | A/T | 0.98 | 1.32 | 0.08 | 1.30e-6 | LOC102723561 |
| 4 | rs138362425 | 75121348 | C/A | 0.98 | 1.35 | 0.09 | 1.37e-6 | MTHFD2L |
| 13 | rs7491779 | 82485534 | A/G | 0.68 | 0.93 | 0.01 | 2.21e-6 | None |

Chromosome (chr), single-nucleotide polymorphism (SNP), base pair (BP), alleles (A1/A2; effect allele/non effect allele), effect allele frequency (F(A1)), effect allele odds ratio (OR), standard error (s.e.).

### Within-Family Polygenic Score sensitivity models

The within-family major depression (MD) polygenic score (PGS) prediction of lifetime MDD described in the main text was repeated in a series of sensitivity analyses. First, data were restricted to sex-concordant dizygotic (DZ) twin pairs (N = 1408; 704 pairs) to evaluate a potentially confounding effect of sex in the main model. The MD PGS significantly predicted lifetime MDD in the global model (OR = 1.434, *p* = 9.52 × 10^-9^), and so did the between- and within-family MDD PGS predictors in the within-family model (OR = 1.409, *p* = 9.04 × 10^-5^; OR = 1.948, *p* = 1.09 × 10^-6^). Unlike the main model, the difference between the between- and within-family MD PGS effects was no longer significant (Χ^2^ (1, *N =* 1408) = 0.609, *p* = 0.435), suggesting sex might be the main confounder in this analysis. Next, data were instead restricted to sex-discordant DZ twin pairs (N = 874; 437 pairs). Here, the MD PGS significantly predicted lifetime MDD in the global model (OR = 1.420, *p* = 3.73 × 10^-4^), and the between- and within-family MD PGS significantly predicted lifetime MDD in the within-family model (OR = 1.291, *p* = 0.039; OR = 2.254, *p* = 1.61 × 10^-4^). Similar to the main model, the difference in the between- and within-family MD PGS effects remained significant (Χ^2^ (1, *N =* 874) = 5.033, *p* = 0.025). Finally, data were restricted to DZ twin pairs genotyped on the same array (N = 2084; 1042 pairs), to evaluate a potential confounding effect of genotype array in the main model. The MD PGS significantly predicted lifetime MDD in the global model (OR = 1.467, *p* = 5.62 × 10^-9^), and so did the between- and within-family MD PGS predictors in the within-family model (OR = 1.447, *p* = 3.43 × 10^-5^; OR = 1.880, *p* = 6.88 × 10^-6^). Unlike the main model, the difference in the between- and within-family MD PGS effects was not significant (Χ^2^ (1, *N =* 2084) = 2.48, *p* = 0.116). The sensitivity analyses suggest that sex differences might be a potential source of confounding of the association between the MD PGS and lifetime MDD.

### **Supplementary Figure 4.** Number of twins by MD PGS decile for discordantly affected and concordantly unaffected pairs.

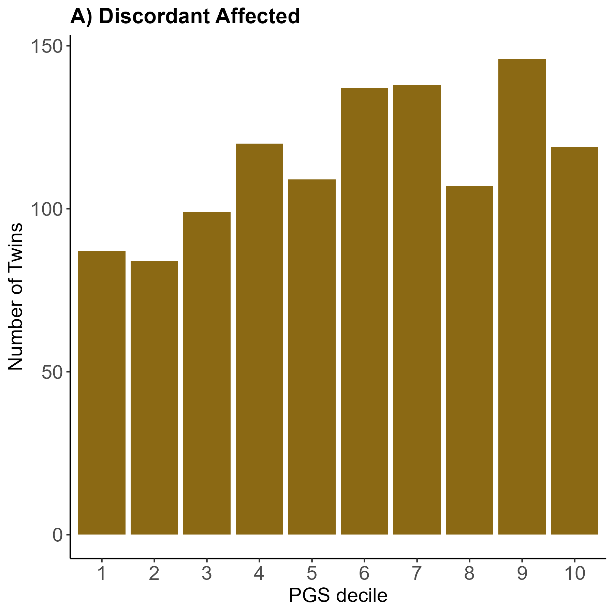

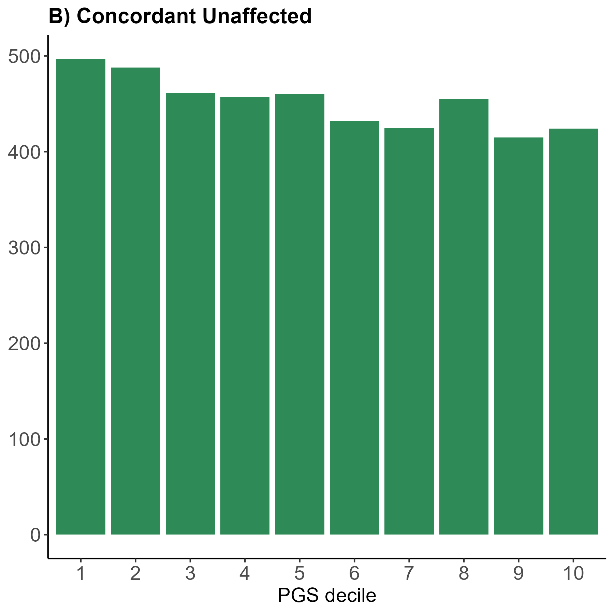

Number of twins per major depression (MD) polygenic score (PGS) decile by monozygotic and dizygotic twin major depressive disorder (MDD) concordance. A) Discordant affected twins (N = 1146). B) Concordant unaffected twins (N = 4514).

### Twin concordance and PGS quartile in the Australian Genetics of Depression Study and NTR

The comparison of twin concordance and MD PGS was assessed in an independent twin sample, the Australian Genetics of Depression Study (AGDS). AGDS is an ongoing study on the genetic determinants of depression in Australia, spanning over 20 000 individuals of which over 15 000 also provided genotype information. PGSs were created using SBayesR software^67^ with weights derived from Adams et al.^68^. We computed MD PGS quartiles in the AGDS genotype sample (sample size did not allow for deciles) and restricted the sample to complete monozygotic (MZ) twin pairs with information on genotype and lifetime MDD status. We then plotted MD PGS quartiles against MZ twin concordance status: concordantly affected (N = 152 pairs), concordantly unaffected (853 pairs), and discordantly affected twins (426 pairs). We repeated the Netherlands Twin Register (NTR) analyses following this approach. Results in AGDS mirrored those from the main analyses, in that the fraction of concordant unaffected twins decreases in higher MD PGS quartiles, and the fraction of concordant affected twins increases in higher MD PGS quartiles (Supplementary Figure 6). This same pattern was observed for the alternative NTR definition (Supplementary Figure 7).

### **Supplementary Figure 5.** Number of twins by MDD PGS quartile and by twin pair MDD concordance in AGDS.

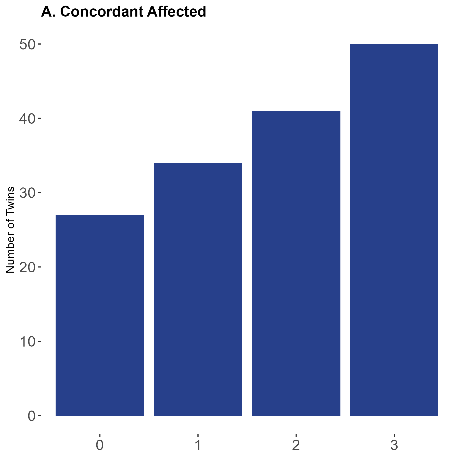

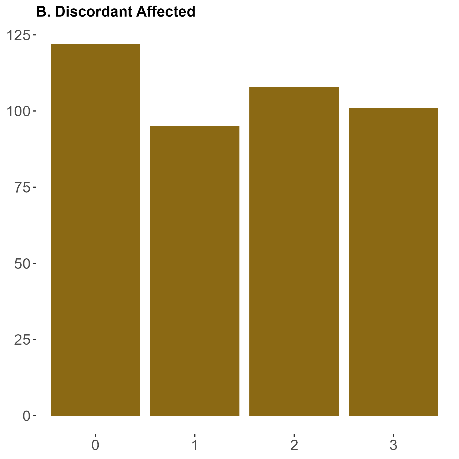

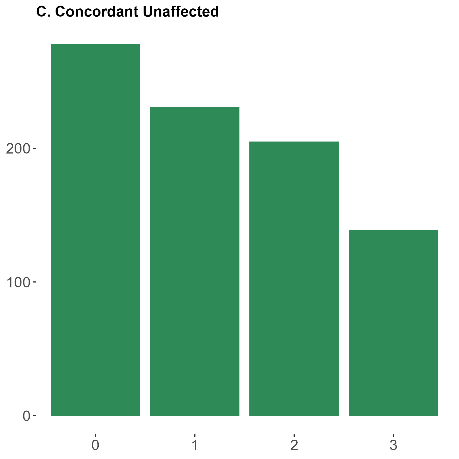

Number of twins per major depression (MD) polygenic score (PGS) decile by monozygotic twin major depressive disorder (MDD) concordance status in the Australian Genetics of Depression Study (AGDS).

### **Supplementary Figure 6.** Number of twins by MDD PGS quartile and by twin pair MDD concordance in NTR.

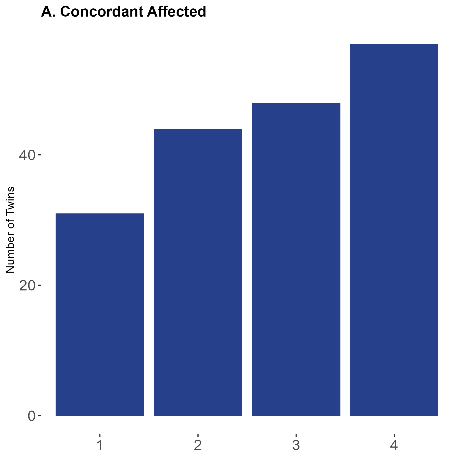

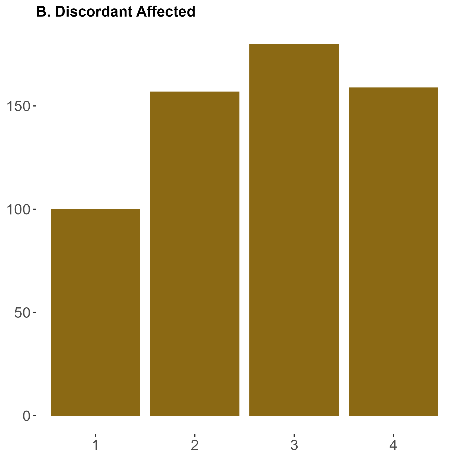

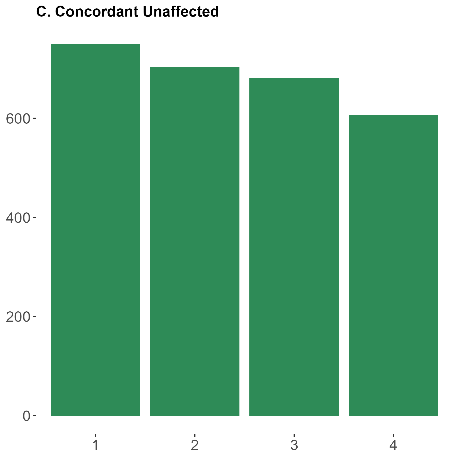

Number of twins per major depression (MD) polygenic score (PGS) decile by monozygotic twin major depressive disorder (MDD) concordance status in the Netherlands Twin Register (NTR).

### **Supplementary Figure 7.** MAGMA tissue enrichment analysis of MDD.

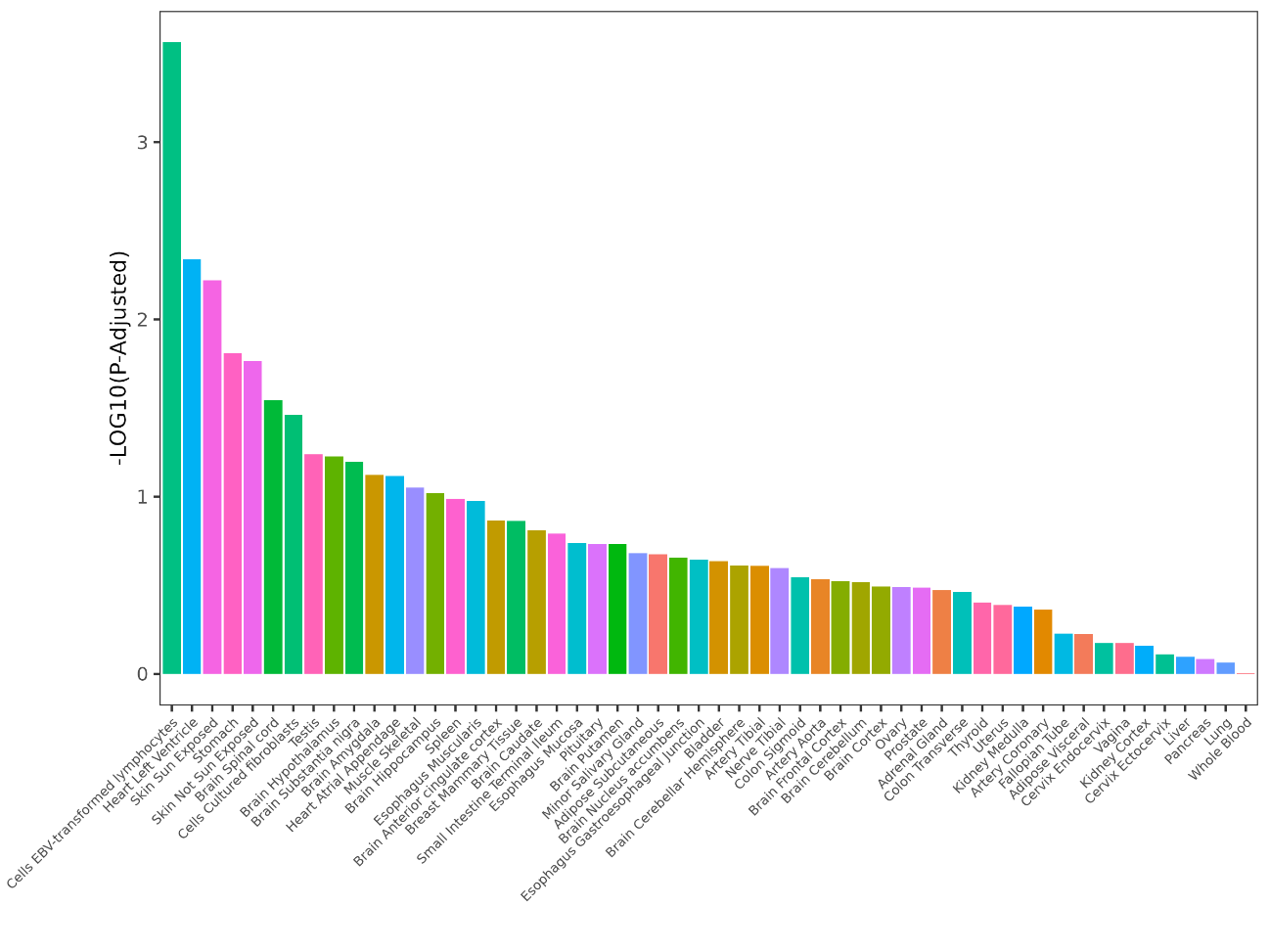

MAGMA tissue enrichment analysis of major depressive disorder (MDD). The x-axis shows 53 tissue types from the GTEx v8 database. The y-axis shows nominal -log10 P-values.

### **Supplementary Figure 8.** Genome-wide association mega-analysis of height.

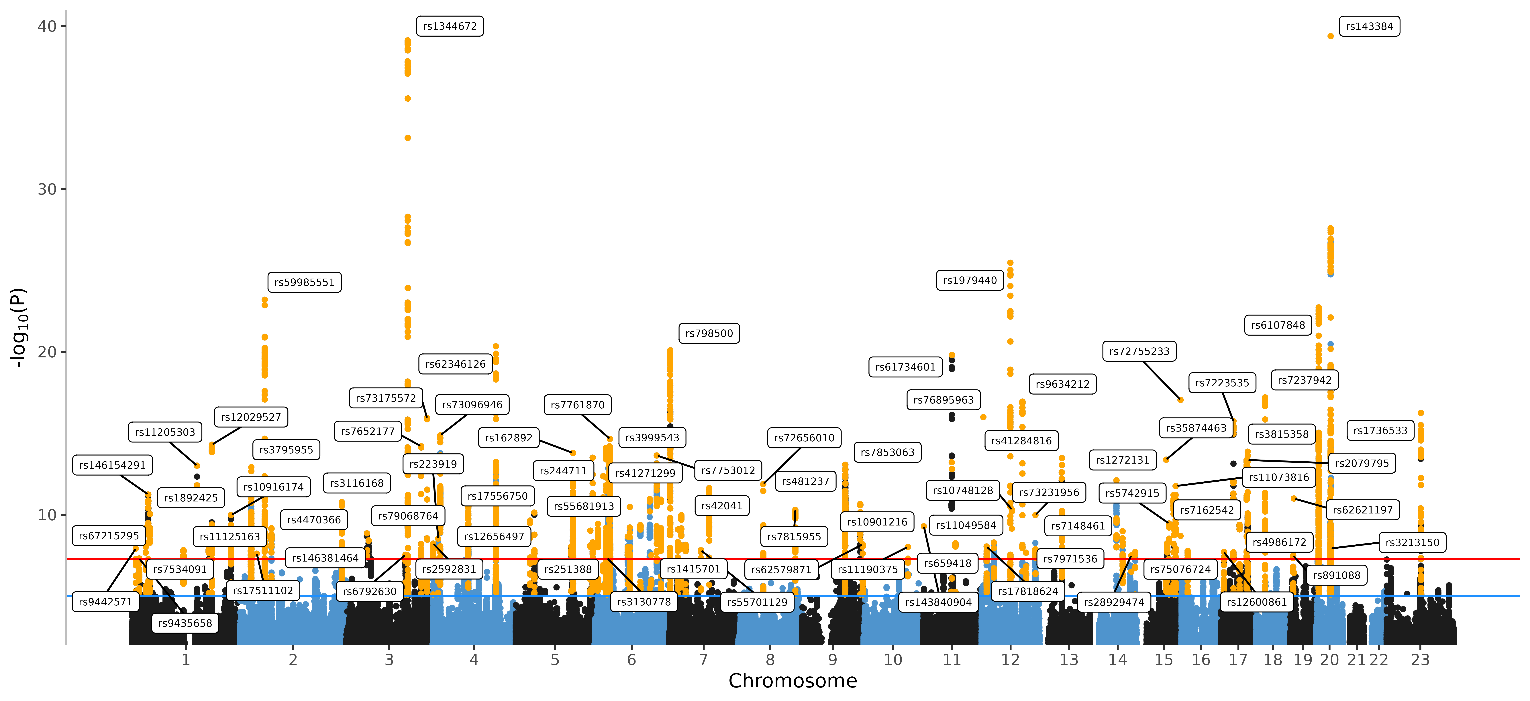

Manhattan Plot of the genome-wide association mega-analysis of height in BIONIC (N = 52 893). The x-axis indicates chromosomal position. Y-axis denotes -log10 *p* and indicates the strength of association between the SNP and outcome phenotype. The red line indicates Bonferroni-corrected genome-wide significance; *p* < 5 × 10^-8^. The blue line indicates suggestive singificance; *p* < 5 × 10^-5^.

### **Supplementary Figure 9.** QQplot of genome-wide association mega-analysis of height.

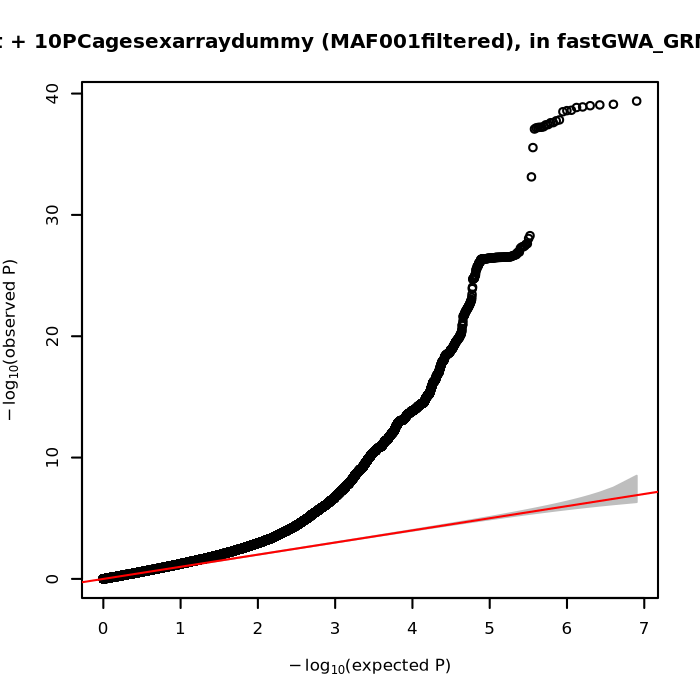

QQplot showing expected versus observed -log10 P-values from the genome-wide association mega-analysis of height (N = 52 893).

### Genome-wide association meta-analysis of MDD

Results from the meta-analysis of MDD are presented in Supplementary Figure 9. The significant finding on chromosome 1 that was found in the GWAS mega-analysis did not reach significance in the meta-analysis, although it remained the top SNP and very close to the significance threshold. LDSC SNP-based heritability (SNP-*h*²) on the liability scale (N_eff_ = 63 743) was SNP-*h*² = 0.134 (0.016). The LDSC genetic correlation (r_G_) with the GWAS mega-analysis (same sample, different method) was *r*_G_ = 1.085 (0.013). The LDSC genetic correlation with the PGC-MD sumstats was *r*_G_ = 0.866 (0.048). Altogether, these results indicate the two methodologies yield highly similar results and are in line with minimal confounding due to genotype array.

### **Supplementary Figure 10.** GWAS meta-analysis of lifetime major depressive disorder case-control status.

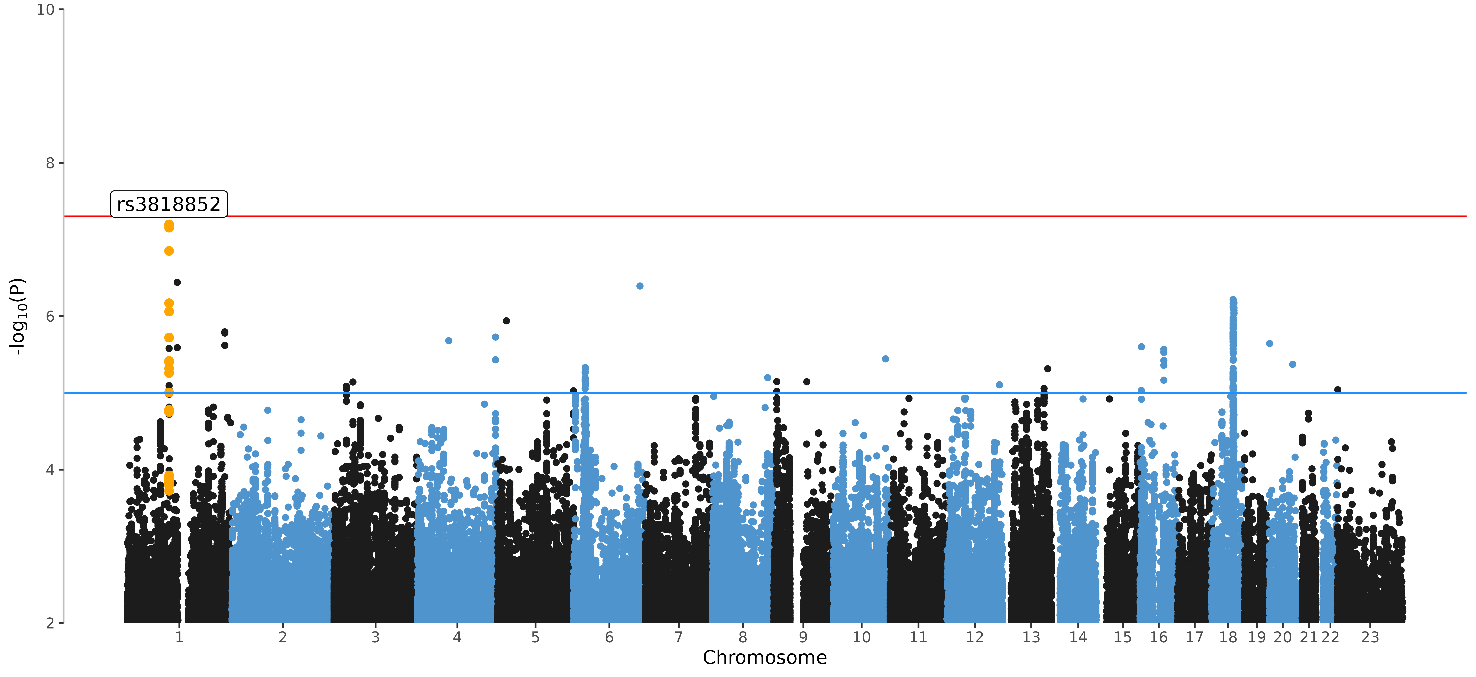

Manhattan Plot of the genome-wide association (GWAS) meta-analysis of lifetime major depressive disorder (MDD) in BIONIC. The x-axis indicates chromosomal position. Y-axis denotes -log10 P-value and indicates the strength of association between the single-nucleotide polymorphism and the outcome phenotype. The red line indicates Bonferroni-corrected genome-wide significance; *p* < 5x10^-8^. The blue line indicates suggestive singificance; *p* < 5x10^-5^.

### Additional screening of lifetime MDD controls

In this model with no additional screening of lifetime MDD controls, we found a SNP-*h*² on the liability scale (N_eff_ = 66 124) of 0.118 (0.015). Genetic correlation with the main model was *r*_G_ = 0.999 (0.001). Genetic correlation with the PGC-MD GWAS results was *r*_G_ = 0.869 (0.050). These results suggest minimal bias due to the exclusion of individuals diagnosed with other psychiatric disorder from the control population.
